# Impact of COVID-19 pandemic on weight and BMI among UK adults: a longitudinal analysis of data from the HEBECO study

**DOI:** 10.1101/2021.07.10.21259585

**Authors:** Samuel J. Dicken, John J. Mitchell, Jessica Newberry Le Vay, Emma Beard, Dimitra Kale, Aleksandra Herbec, Lion Shahab

## Abstract

**Background:** COVID-19-related restrictions impacted weight and weight-related factors during the initial months of the pandemic. However, longitudinal analyses are scarce.

**Methods:** An online, longitudinal study was conducted among self-selected UK adults (n=1,818), involving three surveys during 2020 (May-June, August-September, November-December), covering height, weight and sociodemographic, COVID-19-related and behavioural measures. Data were analysed using generalised estimating equations.

**Results:** Self-reported average weight and body mass index (BMI) significantly increased from May-June to August-September (74.95kg to 75.33kg, 26.22kg/m^2^ to 26.36kg/m^2^, both p<0.001), and then significantly decreased to November-December (to 75.06kg, 26.27kg/m^2^, both p<0.01), comparable to May-June levels (p=0.274/0.204). However, there was great interindividual variation, with 37.0%/26.7% reporting an increase and 34.5%/26.3% reporting a decrease in weight/BMI greater than 0.5kg/0.5kg/m^2^, respectively from May-June to November-December. The average weight/BMI increase was 3.64kg (95% confidence interval: 3.32,3.97)/1.64kg/m^2^ (1.49,1.79), and the average weight/BMI decrease was 3.59kg (3.34,3.85)/1.53kg/m^2^ (1.42,1.63). In fully adjusted models, increase in weight/BMI across surveys was significantly negatively associated with initial BMI, and positively associated with monthly high fat, salt and sugar (HFSS) snacks intake and alcohol consumption, and for BMI only, older age. However, associations were time-varying, such that lower initial BMI, higher HFSS snacks intake and high-risk alcohol consumption were associated with maintenance of increased weight/BMI from August-September to November-December.

**Conclusion:** The average weight/BMI of UK adults increased during the early pandemic months, before returning to baseline levels in November-December 2020. However, this masks substantial interindividual variation in weight/BMI trajectories, indicating vulnerabilities associated with changes in food and alcohol consumption throughout the pandemic.

**What is currently known from previous studies:**

- Small increases in average weight/BMI have been reported (1.57kg/0.31kg/m^2^) during the initial three months of the pandemic.
- Changes in weight/BMI during the early months were not uniform, with significant proportions increasing (11.2-72.4%) as well as decreasing (7.2-51.4%) weight/BMI.
- Weight/BMI change has been associated with several sociodemographic, lifestyle and behavioural factors.
- Whether these weight/BMI changes persist over longer durations of the pandemic, and the factors associated with any long-term weight/BMI change is largely unknown.

**What this paper adds:**

- In UK adults, average weight/BMI first increased and then decreased from May to December 2020 during the pandemic, but this masks large interindividual variability in average changes.
- Initial BMI at the start of the pandemic and health behavioural factors such as alcohol consumption and high fat, salt and sugar (HFSS) snacks intake were significantly associated with a change in weight/BMI.
- The strength of the association of alcohol consumption, initial BMI and HFSS snacks intake with weight/BMI change was dependent on the stage of the pandemic, with more pronounced differences becoming apparent during the latter part of 2020.

## 1. Introduction

Since the start of 2020, the COVID-19 pandemic has seen the introduction of severe lockdown restrictions to limit avoidable morbidity and mortality. In the UK, COVID-19 lockdown restrictions were first imposed on 23^rd^ March 2020. Restrictions were then eased from June and July onwards, before harsher restrictions returned at the start of October. Various forms of lockdowns were then enforced across the four nations of the UK in November and December 2020. The timing and duration of lockdowns varied across devolved nations. All nations were under lockdown for several weeks during November and December before a tiered restriction system was introduced, where some regions remained under strict lockdown whilst other regions faced more relaxed restrictions.

Obesity-related diseases are a major cause of global morbidity and mortality [1]. Within the UK, the prevalence of overweight and obesity stands at over 60% and continues to rise [2]. Health behaviours including physical activity, diet, smoking and alcohol consumption can impact on bodyweight [3–6], and such health behaviours have been shown to be affected by COVID-19 lockdown restrictions [7–10]. Lockdown restrictions can also impact on other individual, social and environmental factors that influence energy intake and expenditure [7,8]. Indeed, reports from the initial months of the pandemic suggest that average weight and body mass index (BMI) have increased significantly by 1.57kg (95% confidence interval: 1.01-2.14) and 0.31kg/m^2^ (95%CI: 0.17-0.45) compared to before the pandemic [11]. However, reports also suggest that despite significant proportions of people increasing weight and BMI (11.2-72.4%), many have also decreased weight and BMI (7.2-51.4%) during the start of the pandemic [11–17]. These initial weight trajectories during COVID-19 are associated with several socio-demographic, COVID-19-related and behavioural factors including age, gender, initial BMI, pandemic living and working conditions, diet, physical activity and alcohol intake [7,9,10,12,18–21]. Short-term shifts in health behaviours can result in small, yet meaningful changes in bodyweight, as seen with seasonal holiday weight gain during the winter months, which accounts for a large proportion of annual weight gain [22,23]. If these weight/BMI changes seen during the COVID-19 pandemic persist or continue to shift with changes in lockdown restrictions, these changes in health risk could have long-lasting impacts on population health.

However, studies to date assessing the influence of the COVID-19 pandemic on weight/BMI have largely been cross-sectional and during the initial months of the pandemic [21]. Other reports have shown that a range of health behaviours and weight-related factors have been impacted over longer periods of the pandemic [12,24,25]. Little is known about the weight/BMI trajectories of individuals beyond summer 2020, nor over longer periods of the pandemic. In the UK, no studies to date compare the strict lockdown conditions of May-June 2020, to the eased restrictions in August-September, and again compared this to the tighter restrictions in November-December 2020.

HEalth BEhaviours during the COVID-19 pandemic (HEBECO) is a longitudinal study in the UK assessing the impact of the COVID-19 pandemic on health behaviours, their influences and their outcomes. Using a longitudinal sample, this study sought to answer the following research questions:

RQ1. What was the average (i) weight and (ii) BMI in UK adults at the beginning of the COVID-19 pandemic, and at 3-months and 6-months follow-ups?

RQ2. To what extent were sociodemographic, COVID-19-related and behavioural factors associated with a change in (i) weight and (ii) BMI in UK adults from the beginning of the COVID-19 pandemic to 6-months follow-up?

## 2. Materials and Methods

### 2.1. Study design

This study is a longitudinal analysis of data from an ongoing online study of adults, the HEBECO study (https://osf.io/sbgru/). The study was approved by the Ethics Committee at the UCL Division of Psychology and Language Sciences (CEHP/2020/759). Self-selected participants were recruited through multiple online channels including paid and unpaid advertisements across social media (Facebook, Google and Reddit) and mailing lists of UK universities, charities, local government and networks within Cancer Research UK and Public Health England. The full recruitment strategy is available online (https://osf.io/sbgru/). Participants gave their consent prior to data collection. Data were captured and managed by the REDCap electronic data system at UCL [26,27]. Participants were followed up at 1, 3 and 6 months after their baseline survey via email (except for participants that explicitly opted out), with up to three reminders sent at each follow-up to complete the survey. The surveys used in this analysis span a period of 8 months since the beginning of the pandemic (May to December 2020). The 3-months follow-up survey corresponds to the periods of eased pandemic restrictions during August-September 2020, and the 6-months follow-up survey corresponds to the tighter restrictions during November-December 2020. This analysis uses data from the baseline, 3-months and 6-months follow-up surveys. The study protocol was pre-registered on the Open Science Framework (OSF) before analysis (https://osf.io/pr68k/). Deviations from the pre-registered protocol are described in the Supplementary Materials.

### 2.2. Study sample

The analysis uses data from UK adults (18+) who completed the baseline survey between 5^th^ May and 14^th^ June 2020 (inclusive) and also provided data of interest at the 6-months follow-up survey as a minimum, for the outcome variables outlined below.

The focus of the study is to identify if any weight or BMI change during the COVID-19 pandemic was long-term and has been maintained for 6 months of follow-up, and to identify the predictors of long-term weight or BMI change from baseline to 6-months follow-up. Therefore, to enable calculating BMI, the analytical sample for RQ1 and RQ2 includes as a minimum, all participants who self-reported height and weight at the baseline survey, and at least also self-reported weight at the 6-months follow-up survey. Participants who did not self-report height or weight at the baseline survey, or only at 3-months and 6-months follow-up surveys, or reported only at baseline and 3-months follow-up surveys, were excluded from the analytical sample. Participants who answered, ‘prefer not to say’, or ‘don’t know’ to the weight or height questions were also excluded, as BMI could not be calculated. The supplementary analyses use complete cases only; those participants who self-report weight measures at all three time points and height at the baseline survey.

### 2.3 Survey

The survey covered a range of topics including sociodemographics, anthropometrics, physical and mental health, social isolation experienced during the COVID-19 pandemic and health behaviours including physical activity, diet, alcohol consumption and smoking. The baseline survey retrospectively covered the same topics from before the COVID-19 pandemic (i.e. pre-COVID-19). The baseline survey took around 20 minutes, and follow-up surveys around 15 minutes.

### 2.4. Measures

Full details of outcome and predictor measures can be found in the Supplementary Materials and on OSF (https://osf.io/pr68k/).

#### 2.4.1 Outcomes

Changes in self-reported weight and BMI were the primary and secondary outcomes of interest, respectively. Self-reported anthropometrics are a reliable measure generally; although, individuals tend to overreport height and underreport weight [28].

Change in self-reported weight: Participants were asked at baseline, and at 3- and 6-months follow-up surveys, *‘How much do you weigh? Please try to be as accurate as possible’*. Participants could answer using a drop-down box in 1lb increments (or equivalent metric measure), or answer ‘don’t know’ or ‘prefer not to say’. Weight was analysed in kg as a continuous variable for both RQ1 and RQ2.

Change in self-reported BMI: At baseline, participants were asked *‘What is your height? Please try to be as accurate as possible’*. Participants could answer using a drop-down box in 1-inch increments (or equivalent metric measure), or answer ‘don’t know’ or ‘prefer not to say’. BMI was calculated as weight (kg)/squared height, with height converted into metres (m^2^). BMI was analysed as a continuous variable for both RQ1 and RQ2. In the sensitivity analysis, BMI was also analysed as a categorical variable using World Health Organisation (WHO) BMI categories [29,30].

For RQ2, we computed continuous change scores in self-reported weight and BMI from baseline to 3-months follow-up and baseline to 6-months follow-up surveys. This produced two change scores each for weight and BMI at 3 months and 6 months, which were combined to form two outcome variables, ‘change in self-reported weight’ and ‘change in self-reported BMI’.

#### 2.4.2. Explanatory variables

Sociodemographic variables recorded at baseline included **gender** (female vs all other), **BMI** (continuous), **age** (continuous), **ethnicity** (white vs all other), **occupation and work from home** (categorical: unemployed (which includes retired persons and full-time parents/carers), employed and working from home, or employed and not working from home) and a **socioeconomic score** based on self-reported income, housing tenure and education. Participants were scored from 0-3; 0 if participants had an income <£50,000, lived in unowned housing and had no higher education, or scored 1, 2 or 3 if participants met 1, 2 or all 3 of having an income of ≥£50,000, owning their accommodation (outright/mortgaged) or having completed higher education.

COVID-19-related variables included **living arrangements** (living alone, living with children (with or without adults), or living with adults only) reported at baseline, **isolation status** (total or some isolation vs general or no isolation) reported at each timepoint and **quality of life**, based on self-reported ratings of quality of living, well-being, social and family relationships (a continuous score from 1-5, 1 = poor, 5 = excellent), also reported at each timepoint.

Behavioural variables were time-variant (reported at baseline, 3- and 6-months follow-up surveys) and time invariant (reported at baseline only). Time-invariant variables included high fat, salt and sugar (HFSS) **ΔHFSS snacks**, **ΔHFSS meals** and **Δfruit and vegetable intake change scores**, derived from pre-pandemic dietary intakes retrospectively reported in the baseline survey being deducted from dietary intakes reported at the time of the baseline survey (i.e. at the start of the pandemic), as initial dietary changes have been associated with maintaining pandemic weight gain [12]. The time-varying behavioural predictors included **HFSS snacks intake, HFSS meals intake, fruit and vegetables intake, physical activity** (reduced physical activity vs all other, given the link between reduced physical activity and pandemic weight gain [7,10,21], based on WHO weekly physical activity recommendations of two days per week for strengthening physical activity, and 150 minutes per week for aerobic physical activity [31]), **alcohol consumption** (≤14 weekly units vs >14 weekly units, based on government low-risk drinking recommendations [32]) and **smoking status** (yes vs no). HFSS foods intake, physical activity and alcohol consumption were assessed at each wave using standard measures (see supplementary material). The food item questions are based on previous research study survey questions and derived from Public Health England’s sugar reduction programme definitions as measures relevant for informing health policy [33–35].

### 2.5. Statistical Analysis

Statistical analysis was conducted in SPSS Statistics version 27 (IBM). Significance was defined as p<0.05. The statistical analysis plan was pre-registered on OSF prior to analysis (https://osf.io/pr68k/).

#### 2.5.1. RQ1

We report cross-sectional baseline participant characteristics in RQ1 (weighted characteristics, using age, gender, country of living, ethnicity, education and income based on data from the Office for National Statistics [36] are provided in the Supplementary Materials).

We report the unweighted means with 95% confidence intervals (95% CI) for (i) weight and (ii) BMI at each timepoint. We also report the percentage of the sample increasing or decreasing (i) weight or (ii) BMI between timepoints, where an increase or decrease was defined as more than a 0.5kg or 0.5kg/m^2^ change, respectively, from the reference timepoint (baseline or 3-months follow-up survey). 0.5kg change was used as the cut-off in weight as it has been previously reported as the average seasonal weight gain, which has parallels to the COVID-19 pandemic [22,23]. 0.5kg/m^2^ change was used as the cut-off in BMI as this has been previously used to define an unchanged BMI [37]. Lastly, we reported the mean change (with 95%CI) in (i) weight and (ii) BMI in those increasing or decreasing weight/BMI between timepoints.

The subsequent analyses make use of repeated, related measures over time. Typical regression models assume independence between measures, and analysis of variance methods do not allow for covariates to be included [38]. Generalised estimating equations (GEEs) are a form of generalised linear model that can handle correlated measures, such as within participants. GEE provides population-level, marginal effect estimates of predictor variables on the outcome, rather than examining subject-level average effects, as with linear regression modelling. GEEs can model a range of outcome data distributions by using different link functions and choosing different covariance structures describing the correlation between repeated measures over time, making the model highly versatile for longitudinal analyses [38]. In this study, we used an unadjusted GEE model to examine the changes in self-reported (i) weight and (ii) BMI across 6 months of follow-up during the COVID-19 pandemic, using the identity link function for a linear scale response, as change in (i) weight and (ii) BMI were normally distributed continuous outcome variables, and the AR(1) covariance structure (given that participant measures at closer timepoints are expected to be more correlated [39]). Pairwise time comparisons were conducted between baseline, 3- and 6-months timepoints. Comparisons were inserted into the GEE model as factors, with multiple comparisons adjusted for using sequential Šidák correction.

#### 2.5.2. RQ2

GEE models were used to determine the association between the sociodemographic, COVID-19-related and behavioural predictor variables listed above and change in (i) weight and (ii) BMI across timepoints. Time*explanatory variable interactions were inserted into GEE models to assess temporal differences in the association of explanatory variables with continuous changes in (i) weight and (ii) BMI over time. The time variable was categorical, as the trajectory of change in weight and BMI was not expected to be linear [40].

An unadjusted GEE model was computed showing the association between each individual explanatory variable and the change in (i) weight or (ii) BMI across timepoints. Each individual explanatory variable model was then adjusted for a main effect of time and for a time*explanatory variable interaction. A fully adjusted GEE model containing all explanatory variables was then computed. The fully adjusted GEE model was assessed for goodness of fit using Quasi-Likelihood under Independence Model Criterion (QIC), which is an appropriate measure of goodness of fit for models using the AR(1) covariance structure [38]. Then, a fully adjusted GEE model containing all explanatory variables as well as all significant time*explanatory variable interactions from the univariate models with time interactions was computed. Time*explanatory variable interactions were retained in the full GEE model if they improved QIC >2 over the full GEE model without interactions, and the interaction itself remained significant (p<0.05).

Independent variables were retained after checking for collinearity using Pearson correlations (all correlations *r*<0.4).

#### 2.5.3. Sensitivity analyses

RQ2 was repeated using complete cases. Further analyses were also conducted using two binary logistic GEE models with the binary logistic model and logit link function for an ‘increase’ in weight/BMI vs ‘all other’, and a ‘decrease’ in weight/BMI vs ‘all other’, with a minimum 5% weight/BMI change from baseline cut-off to define an increase or decrease in weight/BMI. Such magnitude weight change has been used to define clinically meaningful weight loss [41,42].

For BMI only, analyses were stratified by WHO BMI categories [30], i.e. associations with an ‘increased BMI’, vs ‘all other’ were assessed in individuals with a baseline BMI <25kg/m^2^ (normal weight and underweight), and associations with a ‘decreased BMI’ vs ‘all other’ in individuals with a baseline BMI ≥25kg/m^2^ (overweight and obesity).

Further details of this analysis is provided in the Supplementary Materials.

Bayes factors can help with the interpretation of potentially non-significant effects if found during analyses, which is not possible using frequentist statistics [43]. Bayes Factor analyses in the event of non-significant findings were pre-registered online for physical activity, BMI, snacking, alcohol consumption, age and gender (https://osf.io/pr68k/), as current literature suggests that reduced physical activity, higher initial BMI, increased snacking, high alcohol consumption, younger age and female gender are associated with weight/BMI gain at the start of the pandemic [7,8,10,21]. To differentiate non-significant associations from data supporting the null, data supporting the alternative, and inconclusive data, effect sizes were obtained from mean differences in weight change reported in the COVID-19 literature [44]. Alternative hypotheses were modelled using a half-normal distribution with a peak at zero, given that smaller effect sizes nearer to the null are more likely than larger effect sizes [44,45]. The standard deviation (SD) was set to the expected mean difference in weight change reported in the COVID-19 literature for gender and physical activity: a prior mean difference between males and females of 0.20kg weight change during the pandemic [10] and a prior mean difference between maintaining physical activity and reduced physical activity of 0.23kg weight change during the pandemic [10]. Bayes factors were then calculated using an online calculator: http://bayesfactor.info/.

## 3. Results

Out of a total of 2992 UK participants over the age of 18 recruited into the HEBECO baseline survey, 1,818 (weighted = 1,631) participants met the inclusion criteria for analyses. Table 1 shows the unweighted baseline characteristics for the total, included and excluded samples (for weighted participant characteristics, see Supplementary table S1). There were some differences between included and excluded samples. Included participants were more likely to be female, older, of white ethnicity, under stricter lockdown conditions, a non-smoker, unemployed (which includes students or retired persons) and to have a higher baseline BMI, a higher quality of life score and higher socioeconomic score, and consumed more portions of fruit and vegetables and fewer portions of HFSS meals at baseline.

**Table 1.** Unweighted baseline participant characteristics for the total, included and excluded samples.

|  | Total Sample | Included sample | Excluded sample |  |
| --- | --- | --- | --- | --- |
|  | Unweighted (%) | Unweighted (%) | Unweighted (%) | p |
| **N** | 2992 | 1818 | 1174 |  |
| **Gender** |  |  |  | 0.107 |
| All other | 31.4% | 30.3% | 33.0% |  |
| Female | 68.6% | 69.7% | 67.0% |  |
| **Ethnicity** |  |  |  | <0.001 |
| All other | 6.3% | 4.5% | 9.1% |  |
| White | 93.7% | 95.5% | 90.9% |  |
| **Mean BMI [SD] (N=2783)** | 26.1 [5.2] | 26.2 [5.1] | 25.7 [5.3] | 0.018 |
| **Mean Age [SD]** | 47.9 [15.5] | 51.7 [14.3] | 42.0 [15.4] | <0.001 |
| **Occupation and work from home (N=2790)** |  |  |  | <0.001 |
| Unemployed (including retired persons and full-time parents/carers) | 28.6% | 33.2% | 21.5% |  |
| Employed working from home | 51.6% | 48.0% | 57.1% |  |
| Employed not working from home | 19.8% | 18.8% | 21.3% |  |
| **Socioeconomic score** |  |  |  | <0.001 |
| Income <£50k, unowned Housing and no higher education | 4.9% | 4.1% | 6.1% |  |
| 1 of: ≥£50K income, housing ownership/mortgage or higher education | 27.8% | 24.2% | 33.3% |  |
| 2 of: ≥£50K income, housing ownership/mortgage or higher education | 38.5% | 40.7% | 35.0% |  |
| All of: ≥£50K income, housing ownership/mortgage and higher education | 28.8% | 31.0% | 25.5% |  |
| **Living conditions** |  |  |  | <0.001 |
| Alone | 16.8% | 16.8% | 16.9% |  |
| With children (with or without adults) | 19.5% | 17.1% | 23.3% |  |
| With adults only | 63.6% | 66.1% | 59.8% |  |
| **Isolation status (N=2946)** |  |  |  | **0.285** |
| Total or some isolation | 79.3% | 79.9% | 78.3% |  |
| General or no isolation | 20.7% | 20.1% | 21.7% |  |
| **Mean Quality of Life [SD] (1-5) (N=2889)** | 3.4 [0.8] | 3.4 [0.8] | 3.3 [0.8] | **<0.001** |
| **Mean HFSS snacks (portions per month) [SD] (N=2609)** | 58.4 [45.2] | 56.8 [44.1] | 61.4 [47.0] | **0.012** |
| **Mean HFSS meals (portions per month) [SD] (N=2618)** | 6.6 [8.3] | 5.8 [6.6] | 8.0 [10.6] | **<0.001** |
| **Mean Fruit and vegetables (portions per month) [SD] (N=2647)** | 44.0 [18.0] | 45.4 [17.1] | 41.4 [19.3] | **<0.001** |
| **Mean ΔHFSS snacks change score (portions per month) [SD] (N=2609)** | 8.8 [34.5] | 9.4 [33.3] | 7.9 [36.5] | 0.300 |
| **Mean ΔHFSS meals change score (portions per month) [SD] (N=2618)** | -1.4 [7.9] | -1.3 [7.1] | -1.5 [9.0] | 0.462 |
| **Mean ΔFruit and vegetables change score (portions per month) [SD] (N=2647)** | -0.1 [12.5] | 0.02 [11.6] | -0.4 [13.9] | 0.377 |
| **Physical activity (N=2825)** |  |  |  | **0.002** |
| All other | 72.5% | 70.6% | 75.8% |  |
| Reduced | 27.5% | 29.4% | 24.2% |  |
| **Alcohol consumption (N=2772)** |  |  |  | **0.996** |
| $\leq 14$ weekly units | 81.0% | 81.0% | 80.9% | |
| $>14$ weekly units | 19.0% | 19.0% | 19.1% | |
| **Smoking status** |  |  |  | **<0.001** |
| Yes | 18.6% | 14.0% | 25.7% |  |
| No | 81.4% | 86.0% | 74.3% |  |

*\*P-values are for comparisons between the analytical and excluded samples. 133 (weighted = 141) participants were excluded due to reporting 'don't know' or 'prefer not to say' to the height or weight questions at the baseline or 6-months follow-up surveys. SD: Standard Deviation.*

### 3.1. What was the *average (i) weight and (ii) BMI in UK adults at the beginning of the COVID-19 pandemic and at 3- and 6-month follow-ups during the COVID-19 pandemic?*

Weight and BMI both significantly increased from baseline (May-June 2020) to 3-months follow-up (August-September 2020) (74.95kg to 75.33kg, 26.22kg/m^2^ to 26.36kg/m^2^, both p<0.001) (Figure 1). Weight and BMI then significantly decreased from 3-months follow-up to 6-months follow-up (November-December 2020) (75.33kg to 75.06kg, 26.36kg/m^2^ to 26.27kg/m^2^, both p=0.003). Weight and BMI did not significantly differ from baseline to 6-months follow-up (74.95kg to 75.06kg, 26.22kg/m^2^ to 26.27kg/m^2^, p=0.274/0.204). Complete case analysis did not materially alter these findings (Supplementary Tables S2 and S3).

**Figure 1.**
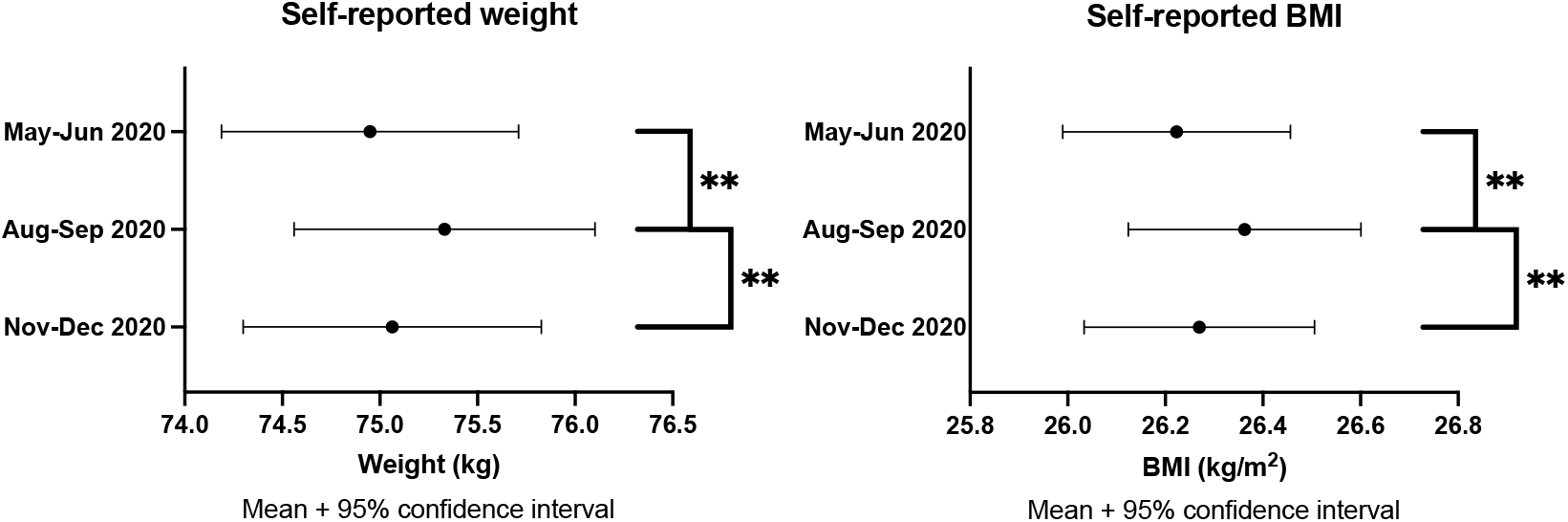
Means and 95% confidence intervals with pairwise comparisons between baseline (May-June 2020), 3 months (August-September 2020) and 6 months follow-up (November-December 2020) for weight and BMI. *\*\*Denotes pairwise comparisons between timepoints were significant at the 0.005 level.*

From baseline, 37.0%/26.7% reported an increase in weight/BMI greater than 0.5kg/kg/m^2^, and 34.5%/26.3% reported a decrease in weight/BMI at the 6-months follow-up survey, compared to the baseline survey (Table 2). Individuals increasing weight/BMI increased on average by 3.64kg [95%CI: 3.32,3.97]/ 1.64kg/m^2^ [1.49,1.79]. Individuals decreasing weight/BMI decreased on average by 3.59kg [3.34,3.85]/ 1.53kg/m^2^ [1.42,1.63].

**Table 2.**
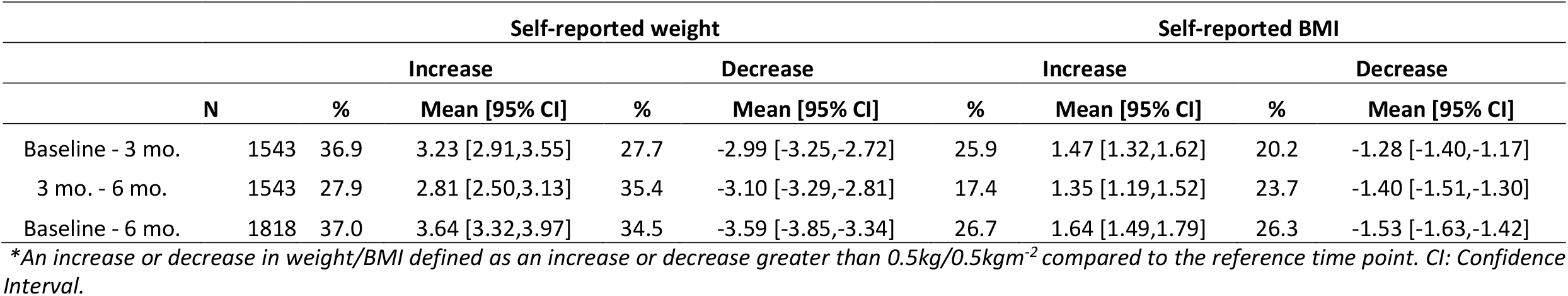
Unweighted proportions increasing or decreasing weight/BMI, and mean changes in proportions increasing or decreasing weight/BMI with 95% confidence intervals.

### 3.2. Which sociodemographic, COVID-19-related and behavioural factors are associated with a change in (i) weight and (ii) BMI in UK adults from the beginning of the COVID-19 pandemic to 6-month follow-up?

In the unadjusted GEE models (Supplementary Table S4), lower baseline BMI, higher HFSS snacks intake, lower fruit and vegetables intake, and a positive (increased intake) ΔHFSS snacks change score were significantly associated with an increase in self-reported weight/BMI. In the fully adjusted GEE model (Table 3), increases in BMI only were associated with older age (B: 0.005kg/m^2^ [<0.001,0.010]). Baseline BMI and HFSS snacks intake remained significantly associated with a change in self-reported weight/BMI, with high-risk alcohol consumption now also significantly associated with an increase in self-reported weight/BMI and neither ΔHFSS snacks change score nor lower fruit and vegetables intake remaining associated. However, based on significant time*explanatory variable interactions from the univariate GEE models (Supplementary Table S4) carried over to the fully adjusted GEE model (Table 3), baseline BMI, HFSS snacks intake and alcohol consumption all demonstrated significant interactions with time and improved model fit. Main effects were not materially altered by the addition of time interactions.

**Table 3.**
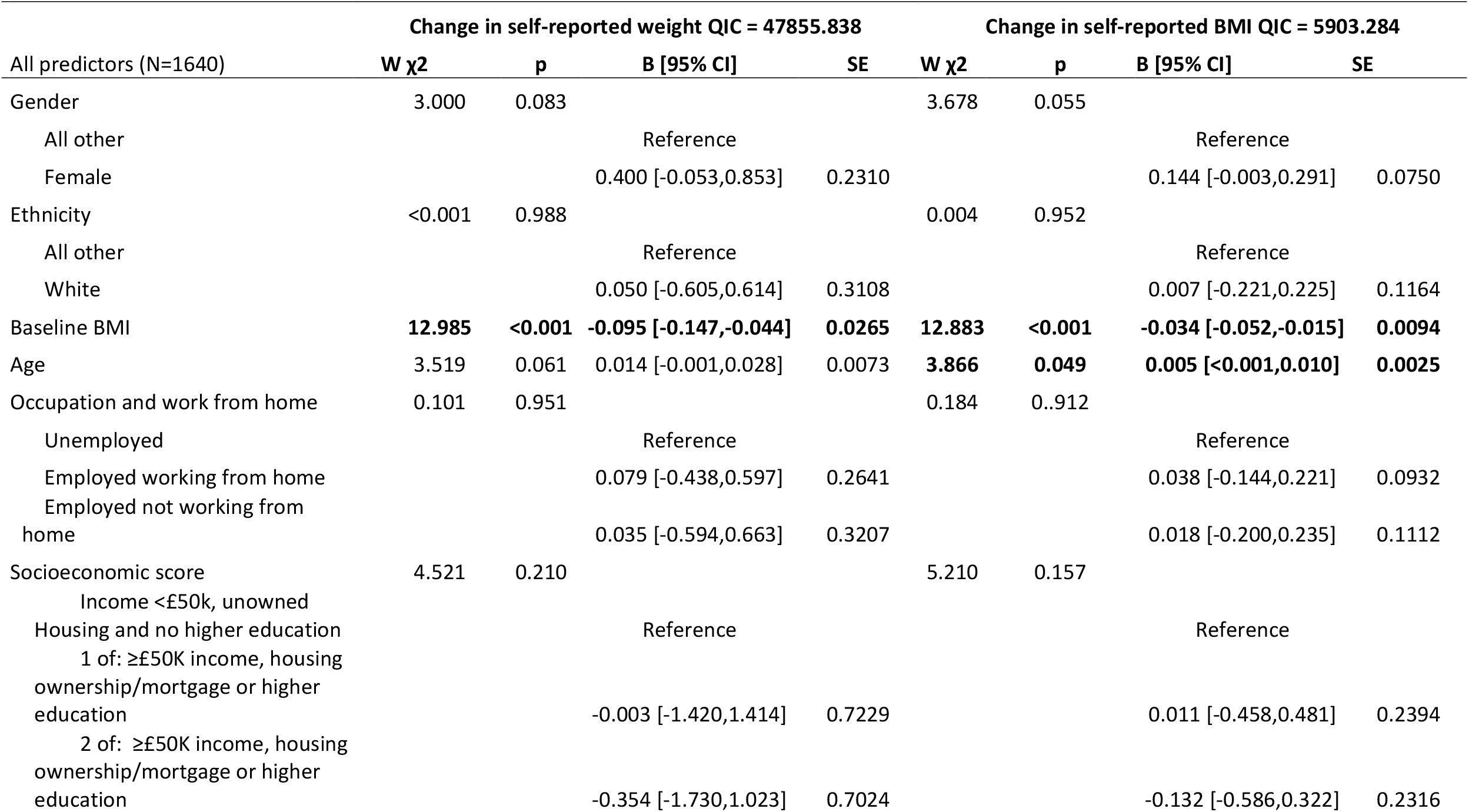
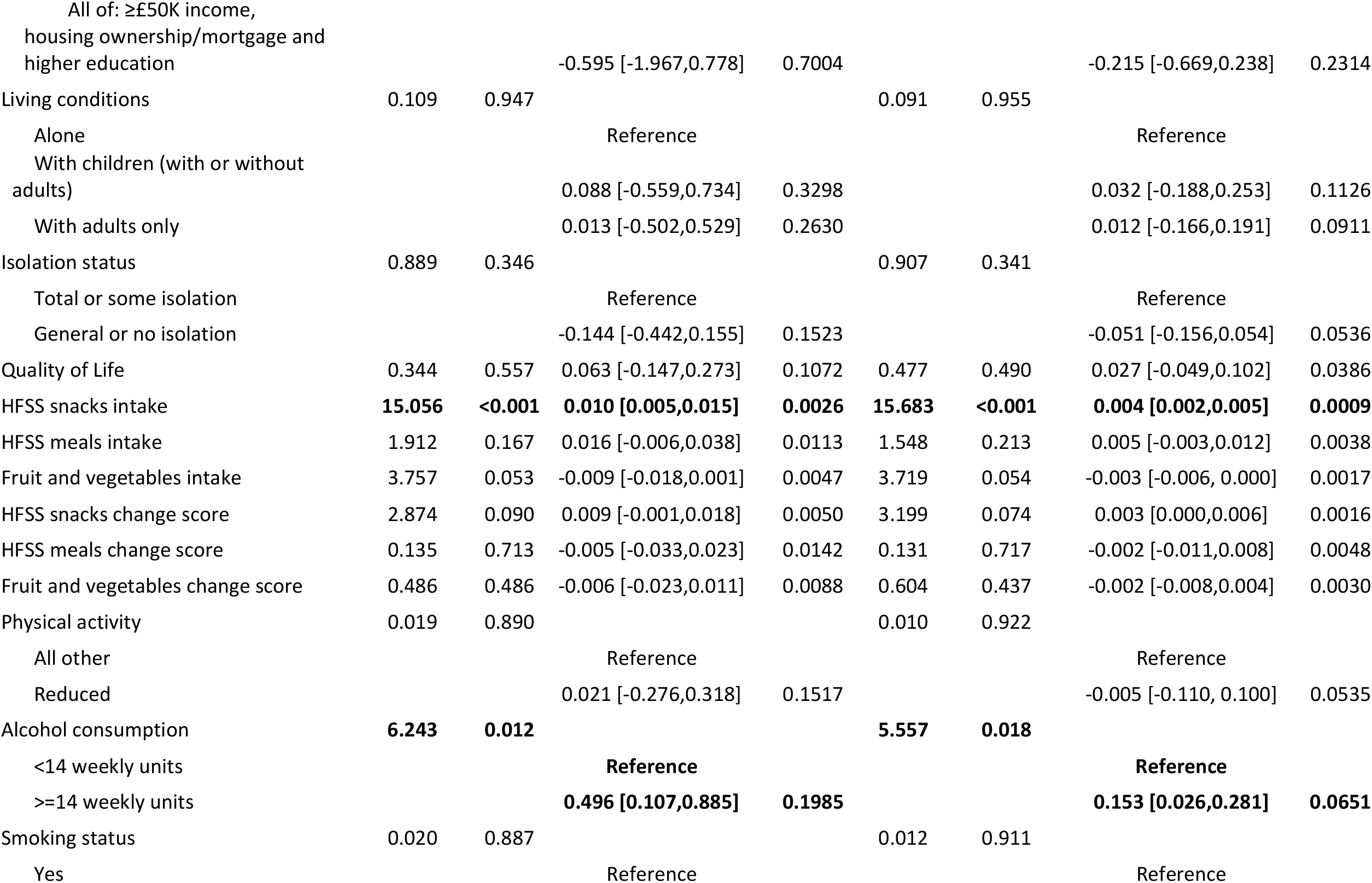
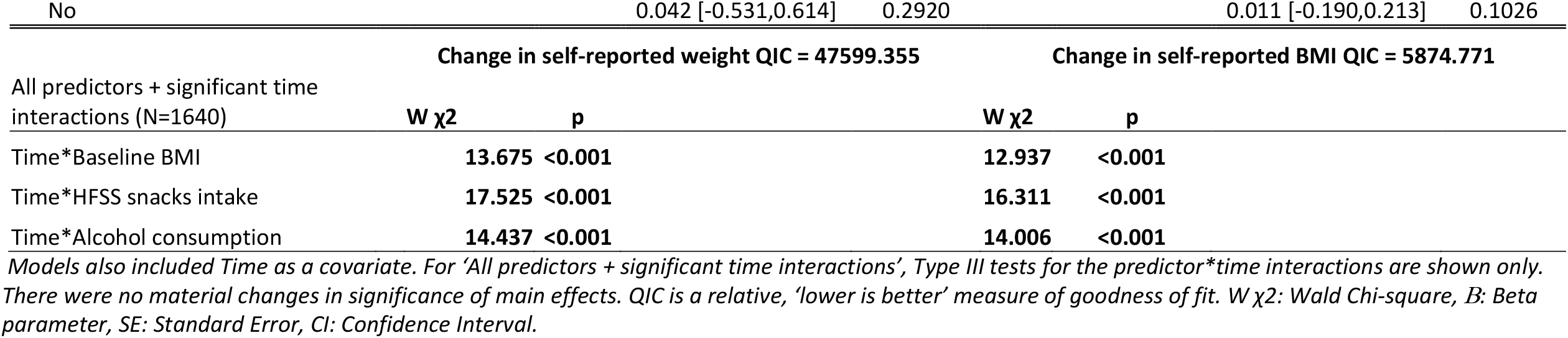
Full GEE model containing all predictor variables and the full GEE model containing all predictor variables and significant predictor*time interactions from univariate models adjusted for time.

Figure 2 schematically demonstrates the time-varying trends of baseline BMI, HFSS snacks intake and alcohol consumption on change in weight/BMI, using a binary cut-off for baseline BMI (<25 vs ≥25) and HFSS snacks intake (below median intake vs median intake and above). The mean weight/BMI change associated with baseline BMI, HFSS snacks intake and alcohol consumption was significantly different at 6-months (November-December 2020) compared to 3-months follow-up (August-September 2020) (all time interactions p<0.001). The association of lower baseline BMI, higher HFSS snacks intake and high-risk alcohol consumption with an increase in weight/BMI was greater at 6-months follow-up than at 3-months follow-up. Pairwise comparisons demonstrated that the change in weight/BMI significantly differed between the binary categories (low-risk vs high-risk alcohol consumption, <25 vs ≥25 BMI, below median vs median and above HFSS snacks intake), at 6-but not at 3-months follow-up.

**Figure 2.**
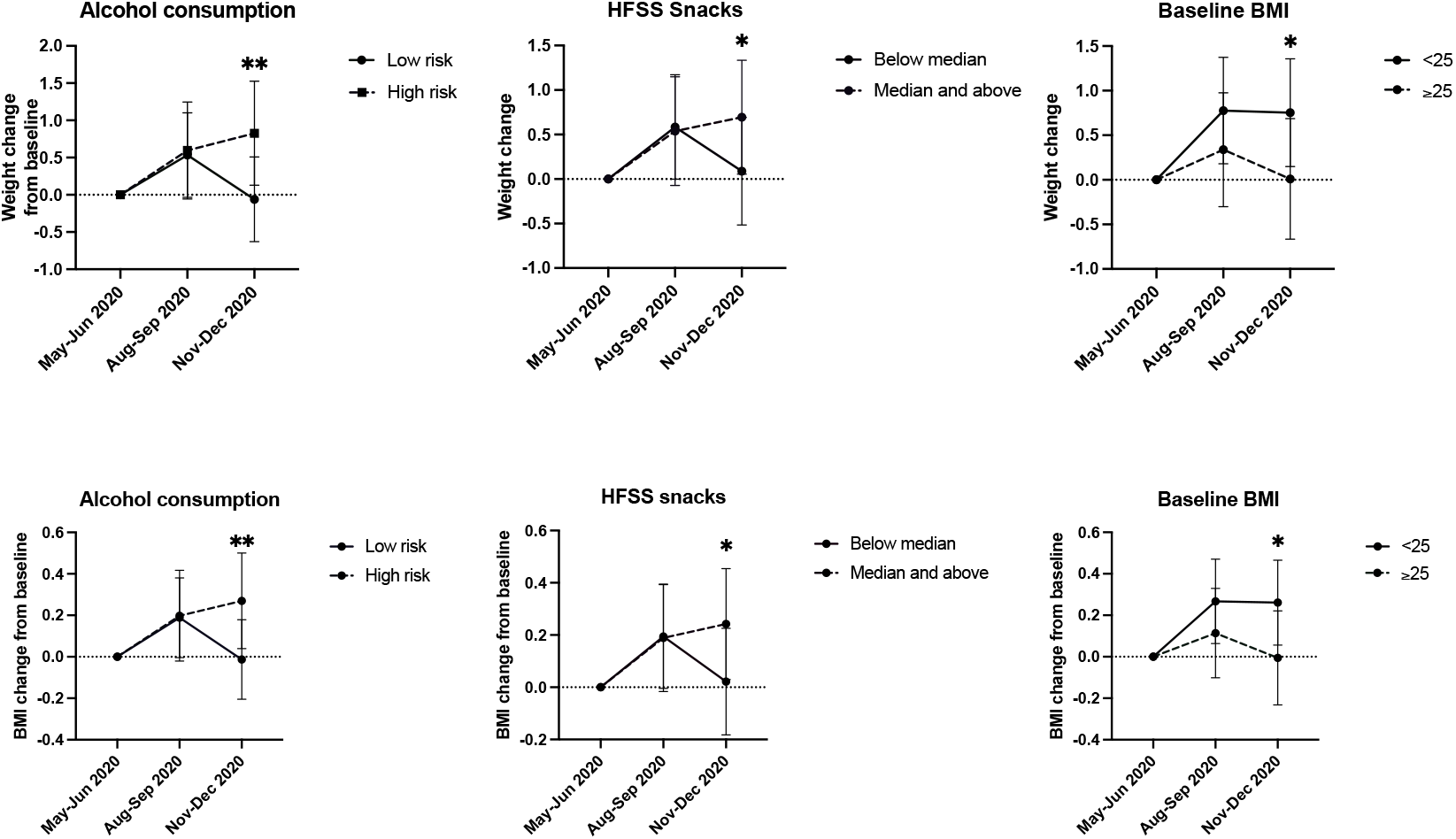
Graphical representations of the time-varying associations of baseline BMI, HFSS snacks intake and alcohol consumption with weight/BMI change at 3-months (August-September 2020) and 6-months follow-up (November-December 2020). *\*Denotes pairwise comparisons between categories were significant at the 0.05 level. **Denotes pairwise
comparisons between categories were significant at the 0.005 level. Median HFSS snacks intake was 34.5 and 40.0 portions per month at 3-months (August-September 2020) and 6-months follow-up (November-December 2020), respectively*

### 3.3. Sensitivity Analyses

Unweighted and weighted proportions of underweight, normal weight, overweight and obesity using complete cases is presented in Supplementary table S5.

In complete case analysis (see Supplementary tables S6 and S7), in the fully adjusted models, baseline BMI and HFSS snacks intake remained significantly associated with a change in weight/BMI. Complete case analysis altered the borderline associations of high-risk alcohol consumption which became non-significantly associated with an increase in weight/BMI and lower fruit and vegetable intake which became significantly associated with an increase in weight/BMI. Age was no longer significantly associated with a change in BMI. Similarly, for both changes in weight and BMI, baseline BMI, HFSS snacks intake and alcohol consumption interactions with time also remained significant and improved the model fit, though main effects were altered slightly by the addition of the time*explanatory variable interactions (Supplementary Table S7).

In the analyses using a binary outcome of a 5% or more ‘increase’ or ‘decrease’ in weight/BMI vs ‘all other’ (see Supplementary Tables S8, S9, S10 and S11), weight increase was associated with a younger age and lower fruit and vegetable intake. Similarly, an increase in BMI in individuals with a BMI <25 was associated with a lower baseline BMI and lower fruit and vegetable intake. Weight decrease was associated with non-female gender, a higher baseline BMI, lower HFSS snacks intake and low-risk alcohol consumption (≤14 weekly units). Likewise, a decrease in BMI in individuals with a BMI ≥25 was associated with non-female gender, younger age, lower HFSS snacks intake, higher fruit and vegetable intake and low-risk alcohol consumption (≤14 weekly units). No time*explanatory variable interactions significantly improved the fully adjusted GEE models for an increase or decrease in weight (Supplementary Table S9), and there were no significant time*explanatory variable interactions for the binary BMI change outcomes (Supplementary Tables S10 and S11).

Because of the null findings reported here for gender and reduced physical activity in the fully adjusted model (Table 3), Bayes Factors were calculated. The Bayes Factors suggest the data were insensitive to detect an effect of gender (BF=2.51) or physical activity (BF=0.60) on change in weight.

## 4. Discussion

In this sample of UK adults, changes in self-reported weight/BMI were seen during the COVID-19 pandemic between May and December 2020. A small but significant increase in average weight/BMI was found at 3-months follow-up (by August-September 2020). However, in the subsequent months of the pandemic, a small but significant decrease in average weight/BMI was observed, resulting in an overall small non-significant increase in average weight/BMI from baseline to 6-months follow-up (November-December 2020). This change, however, does not reflect the considerable interindividual variability in weight/BMI change across the pandemic in 2020. Over half of participants self-reported a change in their weight/BMI at their 6-month follow-up survey compared to their baseline survey at the start of the pandemic. The small, non-significant increase in average weight/BMI at 6-months follow-up in this study appears to reflect a slightly greater proportion of UK adults increasing their weight/BMI than those decreasing their weight/BMI, albeit by similar amounts. A lower baseline BMI, higher HFSS snacks intake and high-risk alcohol consumption were significantly associated with a change in self-reported weight/BMI across the pandemic. Older age was also significantly associated with an increase in self-reported BMI. These associations were time-varying, such that a lower baseline BMI, higher HFSS snacks intake and high-risk alcohol consumption were associated with maintaining the initial weight/BMI increase experienced at 3-months follow-up (August-September 2020) through to 6-months follow-up (November-December 2020). Taken together, our results suggest that diet and alcohol consumption during the pandemic are associated with longer-term changes in weight and BMI. These factors relate to maintaining the initial weight/BMI gain, which is masked by the return to baseline levels, as seen in the population at large.

### Comparison to previous studies

A systematic review and meta-analysis also demonstrated an overall trend for a small, significant increase in population mean weight (1.57kg) and BMI (0.31kg/m^2^) during the initial months of the pandemic [11,12]. A longitudinal study in the US showed that average weight/BMI was still significantly increased above peak-lockdown levels (April-May 2020) by September-October 2020 [12]. However, many reports during COVID-19 show that despite a small increase in population weight/BMI, many individuals have experienced a reduction in weight/BMI [9,10,13–17]. The average weight increase/decrease reported in this study are larger than the 0.6-3.0kg/2.0-2.9kg average weight increase/decrease reported in a systematic review of COVID-19 studies, until July 2020 [7].

Weight changes can be both beneficial and detrimental to health and wellbeing, and understanding the factors associated with these changes is crucial for the development of targeted interventions. Increased HFSS food intake, snacking and alcohol consumption have all been previously identified as important predictors of initial pandemic weight/BMI gain [7,9,10,12,18–21,25]. Studies in the UK and internationally report large inter-individual variability in dietary changes as a result of the COVID-19 pandemic [46], which in turn have been associated with weight change [7,12,14]. Sustained high intakes of ultra-processed, HFSS foods beyond peak lockdown have been associated with maintaining any weight gained during the start of the pandemic [12]. Previous studies have also identified changes in physical activity as an important predictor of weight change [7,9,10,12,18–21], and having an overweight or obese BMI or being female as important predictors of initial pandemic weight gain [7,10,14,47]. In our study, a reduction in physical activity was not significantly associated with a change in weight/BMI, initial BMI was inversely associated with weight/BMI gain, and gender was not significantly associated with a change in weight/BMI in the main analyses between May and December 2020. However, being female was associated with lower odds of decreasing weight/BMI in the sensitivity analyses. Bayes Factor analysis demonstrated that the data in this study was insensitive to detect an effect of gender or physical activity on weight change. One of the few cross-sectional studies beyond the initial months during October 2020 also found no impact of gender on weight gain [25].

The pandemic and its restrictions have resulted in lifestyle shifts for many individuals, ranging from minor to major changes. The early stages of the pandemic have been compared to the holiday season, where short-term shifts in lifestyle can impact on long-term weight management [22]. The pandemic may have resulted in behavioural changes due to social restrictions imposed, changes to working patterns, or from COVID-19-related stress and the coping strategy used [48]. But whether individuals adaptively coped (e.g. through physical activity or fruit and vegetable intake) or maladaptively coped (e.g. through alcohol consumption or HFSS snacking) differs greatly between individuals within studies [49], with maladaptive strategies being linked to pandemic weight gain [12,48]. Individuals may have used maladaptive coping strategies such as increased snacking or alcohol consumption at the start of pandemic, but then sustained these behaviours through habit formation and subsequently maintaining any initial weight/BMI increase after lockdown restrictions were eased.

Dietary changes are important determinants of weight change as brief periods of weight gain are typically driven by energy overconsumption, such as from increased ultra-processed, HFSS food intake [22,47,50,51]. Studies have noted increased snacking alongside elevated stress, appetite, boredom, low craving control and higher emotional eating during the pandemic [12,48,52]. Given the high palatability of HFSS snacks, individuals may have demonstrated emotional eating behaviours and consumed additional HFSS snack foods as a comfort mechanism to deal with COVID-19 related stress, depression or anxiety [52,53]. HFSS foods also tend to have long shelf lives, be cheaper and more readily accessible. Individuals may have ‘stocked-up’ on HFSS foods through less frequent shopping trips [52]. Being stuck at home may have increased sedentary time and time spent watching TV, further contributing to increased snacking [54]. Initial shifts in HFSS snacking behaviours may then have been maintained through habit formation, increasing the risk of energy overconsumption and weight gain.

As with HFSS snacks, alcohol consumption may have been used as a maladaptive coping mechanism for COVID-19-related stress, boredom and depression [55,56]. Consumption may have increased through both a greater frequency of binge-drinking and frequency of consumption. High-risk alcohol consumption has been associated with unfavourable dietary changes during the pandemic [55], possibly by influencing appetite and food choice, thus increasing energy intake both directly and indirectly [54,56]. Alcohol consumption may also impact on sleep quality [56], further contributing to stress and the risk of weight gain [57]. Alcohol consumption may also shifted from the closures of bars and pubs, with more purchases made for home consumption [8].

Reductions in physical activity may not have resulted in weight/BMI change due to simultaneous compensatory behaviours; those exercising more may also have consumed more food, and those exercising less may have consumed less food. There is also a complex relationship between physical activity and alcohol consumption; individuals may have attempted to compensate for high-risk alcohol consumption with additional physical activity in an attempt to offset energy intake [58–60]. Furthermore, the physical activity measure in this analysis assessed strengthening and moderate-vigorous physical activity only and did not consider light physical activity such as walking, other forms of physical activity such as housework or gardening, or forms of physical inactivity, which may have provided a better overall measure of changes in total physical activity and relations to weight change [8,12,21].

The lack of association between female gender and weight/BMI gain in this study may reflect the coping strategies of female participants, and the complex gender differences in changes in health behaviours during the pandemic. Some studies find women increasing exercise more than men [61], being less likely to alter their alcohol intake [58] and being more likely to increase fruit and vegetables intake, eat less, or partake in healthy eating [8,21,61]. However, UK women have also been shown to eat both more and less during the pandemic [18]. Greater COVID-19-related stress has been reported in females during the start of the pandemic, but psychological distress tended to return to pre-pandemic levels by mid-2020 [62,63]. Further, the impact of the COVID-19-related stress on weight/BMI management likely depends on the coping strategies employed, with women in the UK more likely to use any coping strategy compared to men [49].

### Policy implications

Prior to the pandemic, around two thirds of UK adults were living with overweight and obesity, increasing year on year [2]. The results of this study suggest that the pandemic is associated with longer term changes in weight/BMI, which could contribute to the existing obesity epidemic. Considering that obesity alongside adverse dietary choices and alcohol consumption is one of the leading causes of DALYs and years of life lost in the UK [64,65], there is greater urgency than ever to support people to make healthy behavioural choices to support weight management.

The association between high HFSS snacks consumption and weight/BMI gain highlights the need for policy action regarding diet. Greater effort needs to be made to increase the accessibility and availability of healthy dietary choices, and not just limiting the accessibility of unhealthful alternatives.

If maladaptive strategies to stress are resulting in changes in health behaviours that impact on weight management, then greater support for encouraging adaptive coping strategies should be considered. For example, providing resources for individuals to develop coping strategies towards benefitting from the positive mental affect of physical activity, practicing mindfulness, breathing or encouraging sufficient sleep [54]. The associations here further highlight the impact of high-risk alcohol consumption during the pandemic on public health; the fact that persistent high-risk drinking carries with it the risk of maintained weight gain, and the combined risk of higher weight and alcohol on liver function [56]. Additional work needs to be made to screen for individuals with high-risk alcohol consumption, advising on how to avoid using alcohol to deal with stress and anxiety [56], and providing adequate treatment to support a reduction in alcohol intake [66].

### Strengths and limitations

This study has several strengths. This is the first UK study examining weight change across the first year of the COVID-19 pandemic from May to December 2020. This longitudinal study builds and expands upon the largely cross-sectional literature published to date, providing a greater understanding of the long-term impacts of the pandemic on weight management. The analysis included a range of variables that reflect the wide-ranging impact of the pandemic (including sociodemographics, lifestyle, wellbeing, and health behaviours), and with time-varying measures to reflect the changing conditions of the pandemic over time. An important further contribution of this study is that it assessed the associations of a range of health behaviours relevant to weight management (physical activity, diet, alcohol and smoking). The use of GEE modelling for the longitudinal analysis provided several advantages over typical analytical methods. Finally, the use of both weight and BMI as outcome measures, and complete case analyses and sensitivity analyses with binary cut-offs demonstrating largely consistent associations of baseline BMI, HFSS snacks intake and alcohol consumption with weight/BMI management indicates the robustness of these findings.

However, several limitations may have introduced bias. First, the study sample was self-selected, and featured a predominantly female, younger, well-educated cohort. Second, there were differences in ethnic diversity between the included and excluded samples, and data were unweighted for the longitudinal analyses. All of which may limit the generalisability of the findings to the UK population. Third, due to the observational nature of this study, causality cannot be determined. Fourth, as is the case for the majority of epidemiological studies during the COVID-19 pandemic [11], measures of interest were self-reported, including for weight and height that may be underreported and overreported, respectively [28]. It was also not possible to determine whether and how participants measured their height and weight. However, change scores as outcome variables with participants acting as their own controls helped to minimise within-subject measurement error, though it cannot be guaranteed that participants used the same measurement methods across time. Also, participants were not explicitly asked if their weight had changed, nor told that weight change was an outcome of interest, reducing the risk of expectation bias. Previous analyses have shown that longitudinal repeat measures of weight and height over time in older UK adults are highly stable over years of follow-up [67]. While the use of BMI as an outcome carries several limitations [68], at a population level, there is a reliable association with long-term health risk [69]. Fifth, the study did not consider body composition, which could provide further insights into the weight-related health impacts of the pandemic. Sixth, previous studies have compared weight/BMI to pre-pandemic levels, whereas this study compared weight/BMI to baseline levels at the start of the pandemic. Although the baseline survey happened at the beginning of imposed restrictions, given the large weight change from pre-to during-pandemic [11], this may have underestimated the changes in weight/BMI in this study. Lastly, participants were asked about their behaviours in the past week or month, which may have introduced a recall bias.

## 5. Conclusion

Through self-reported survey data, this study in UK adults found a small, but significant increase in weight/BMI during the first three months of the pandemic, and a small, yet significant decrease in weight/BMI during the following three months. Overall, average weight/BMI was not significantly different in November-December 2020 compared to May-June 2020. However, this masks wide interindividual variability in weight/BMI change experienced by many participants in this study. Older age was associated with an increase in BMI, and baseline BMI, and HFSS snacks intake and alcohol consumption showed time-varying associations with increasing both weight and BMI, primarily being associated with maintenance of initial weight gain in early 2020 through to the latter part of 2020. These findings provide detail on the long-term health impacts of the pandemic on weight change, to guide health policy and direct attention to those at increased risk of likely poorer health outcomes associated with weight gain. Further studies should continue to examine the impact of the ongoing pandemic on weight.

## Data Availability

Data are available upon request.

## Ethics

The study was approved by the Ethics Committee at the UCL Division of Psychology and Language Sciences (CEHP/2020/759). Participants gave their consent prior to data collection, and the study was GDPR compliant.

## Author contributions statement

S.D., J.J.M, J.N.L.V, E.B., D.K., A.H., L.S.; conceptualisation and methodology, E.B.; statistical support, S.D., J.J.M.; formal analysis, S.D.; first manuscript draft, L.S., D.K., A.H.; writing, reviewing and editing, A.H., L.S.; supervision. All authors approved the final manuscript.

## Informed Consent Statement

Informed consent was obtained from all subjects involved in the study.

