## Supplementary Materials for "Impact of COVID-19 pandemic on weight and BMI among UK adults: a longitudinal analysis of data from the HEBECO study"

#### 1. Substantive deviations from pre-registered protocol

The AR(1) covariance structure was used for all generalised estimating equation (GEE) models, as the most representative correlation structure between timepoints. The final GEE model was originally to include only the explanatory variables that were significant in three separate GEE models containing sociodemographic, COVID-19-related or behavioural explanatory variables, but was changed to contain all explanatory variables as the aim was to identify significant variables whilst adjusting for all other variables to guide health policy, rather than to build the best predictive model for a change in self-reported weight/BMI. GEE models containing sociodemographic, COVID-19-related and behavioural explanatory variables were therefore redundant and not computed. In the sensitivity analysis for a change in BMI, a change of 5% was used as the cut-off for an 'increase' or 'decrease' in BMI instead of a change in BMI categories because fully adjusted GEE models using the categorical change outcome did not compute due to zero 'all other' ethnicity participants self-reporting a categorical increase or decrease in BMI. The 5% cut-off reflected the 5% cut-off used in the sensitivity analysis for weight.

The pre-registered protocol in RQ1 stated that the proportions of the sample increasing or decreasing (i) weight and (ii) BMI between timepoints and mean change (with 95% confidence intervals) in (i) weight and (ii) BMI (as a continuous variable) in those increasing or decreasing between timepoints would be obtained via an unadjusted, unweighted GEE model. This was instead obtained using simple descriptive statistics.

The alcohol consumption explanatory variable was changed from the pre-registered Alcohol Use Disorder Identification Test for Consumption (AUDIT-C) score to a binary measure of weekly unit frequency due to an error in the survey that did not allow for an AUDIT-C score to be computed at the 3-months follow-up survey. The alcohol variable was changed to high-risk or low-risk alcohol consumption, based on UK government recommendations. The categorical baseline BMI explanatory variable was changed to a continuous baseline BMI predictor variable to increase power and simplify interpretation.

Bayes Factors were calculated for non-significant findings for both weight and BMI outcomes.

#### Bayes Factors Robustness checks

Robustness checks were conducted for Bayes Factors using alternative priors. For gender, a prior mean difference between -0.23kg and 6.1kg produced the same outcome of inconclusive evidence for an effect. For reduced physical activity, a mean difference within -0.390kg and 0.490kg produced the same outcome of inconclusive evidence for an effect for the same standard error.

### Testing linearity of logit in the binary sensitivity analysis models

Linearity of logit in the continuous explanatory variables was tested in the binary sensitivity analysis models for an increase/decrease in weight/BMI vs all other. Continuous explanatory variables were Box-Tidwell transformed. Continuous explanatory variables and transformed variables were then all entered into binary logistic GEE models for an increase/decrease in weight/BMI vs all other. The linearity of logit assumptions were violated in the 'Decrease weight vs all other' model for the baseline BMI, HFSS snacks intake, fruit and vegetable intake, and ∆HFSS snacks change score continuous variables. In the 'Increase BMI vs all other' GEE model, linearity of logit assumptions were violated for the baseline BMI continuous variable. The above variables were replaced where assumptions were violated in univariate and fully adjusted models (Supplementary tables S8, S9, S10 and S11), with the following categorical alternatives:

- Baseline BMI:
  - 'Decrease weight vs all other': two categories based on WHO classifications; <25, ≥25.
  - 'Increase BMI vs all other' (with <25 BMI): two categories based on WHO classifications; underweight, normal weight.
- HFSS snacks intake: the median binary cut-off as used in Figure 2.
- Fruit and vegetable intake: consuming a few portions per day vs all other.
- ∆HFSS snacks change score: increased intake vs all other.

### Detailed description of study explanatory variables

The full wording of questions can be found at: https://osf.io/bja7g/

Time-invariant explanatory variables are obtained from participants' baseline responses. Time-variant explanatory variables are collected at baseline, 3-months and 6-months surveys.

### Time-invariant

### Socio-demographic

- **Gender** (2 levels) *'Which of the following best describes how you think of yourself?'* **female** (female), **all others** (male, non-binary, prefer not to say)
- **BMI at baseline.** Continuous. As described in 2.4.1.
- **Age.** Continuous*. 'What is your age? (years)'.*
- **Ethnicity** (2 levels). *'What is your ethnic group?'* **white** (1 white), **all others** (Mixed/Multiple ethnic groups, Asian/Asian British, Black/African/Caribbean/Black British, Chinese, Arab, Other ethnic group, prefer not to say)

- **Occupation and working from home** (3 levels) **Unemployed, Employed from home, Employed not from home.**
    - All participants were asked, *'What is your current MAIN occupation (during COVID-19)?'* Laid off during COVID-19, unemployed since before COVID-19, retired, homemaker, full-time parent or carer, unable to work due to disability, other, employed (full or part-time), self-employed (full or part-time), student, furloughed during COVID-19.
    - Participants who answered as employed (full or part-time), self-employed (full or part-time), student, furloughed during COVID-19 to the question *'What is your current MAIN occupation (during COVID-19)?'* were asked, *'Can you do your work or study from home?'* Yes, I can do all the work or study from home, I can only do some work or study from home, or No, my work or study cannot be done at home.
    - **Unemployed** (Laid off during COVID-19, Unemployed since before COVID-19, Retired, Homemaker, Full-time parent or carer, Unable to work due to disability, Other), **Employed from home** (Employed AND Yes, I can do all the work or study from home, OR I can only do some work or study from home) **Employed not from home** (Employed AND No, my work or study cannot be done at home).
- **Composite socioeconomic score:** Sum score of education level and combined housing tenure and household income (Sum score 0-3):
    - **Education level.** *'What is the highest level of education that you have completed?'* **0,** below A-level (No formal qualification, GCSE/School certificate/O-level/CSE, Vocational qualifications (e.g. NVQ1+2)), or **1,** A-level or above (A-level/Higher school certificate or equivalent (e.g. NVQ3), Bachelor degree or equivalent (e.g. NVQ4), Masters/PhD/PGCE or equivalent, Other).
    - **Housing and Income.** *'What is your housing tenure?'* Owned outright, Mortgage Rented from local authority, Rented from private landlord, Belongs to housing association, Shared ownership (part owned, part rented), Other. '*What was your household annual income BEFORE COVID- 19?'* up to 13 499 GBP, 13 500-24 999 GBP, 25 000-49 999 GBP, ≥50 000 GBP, prefer not to say. **2** (Housing **=** Owned outright OR mortgage AND Income = 50 000+ GBP), **1** (Housing = All other AND Income = 50 000+ GBP), **1** (Housing = Owned outright OR mortgage AND Income = All other), **0** (Housing = All other AND Income = All other).

**COVID-19 related (impact on well-being and lifestyle)**

- **Living arrangements (3 levels): alone, with children (with or without adults), with adults only.**
    - **Alone.** Participants responding 'I live on my own' when asked *'How many persons other than yourself (including children) LIVE WITH YOU NOW in the same flat or hous*e? Participants answering, 'I live with 1 or more people' were

then asked: *'Do you live with any of these persons below?'* Being able to select multiple responses: With my partner, husband/wife, boyfriend/girlfriend, with children 0-5 years old, with children 6-15 years old, with persons aged 16-69 (family or friends), with persons aged 70+ (family or friends), persons who you believe may be vulnerable to COVID-19 for any reason, persons who are in poor health
  - **With any children (with or without adults)** (if participants respond with 'children 0-5 years old' or 'with children 6-15 years old')
  - **Adults only** (all other participants who did not say 'I live on my own' or did not select 'with children 0-5 years old' or 'with children 6-15 years old')

**Time-variant**

**COVID-19 related (impact on well-being and lifestyle)**

- **Isolation** *'Which type of COVID-19 induced isolation are you experiencing?'* (2 levels) **total and some** (Total isolation/quarantine, Some isolation), **general and no isolation** (General isolation, No isolation)
- **Composite wellbeing, social and living measure**: At baseline: *'How would you rate the below aspects of your life in the since COVID-19?'* At follow-up: *'How would you rate the below aspects of your life in the past month?'* 1-5: 1 poor, 2, 3 average, 4, 5 excellent. Continuous mean score of living + wellbeing + social + family (1-5)

**Behavioural: time-variant**

**Diet:**

- **HFSS snacks intake** continuous
- **HFSS meals intake** continuous
- **Fruit and vegetable intake** continuous
  - Participants were asked at each survey, *'In the past month, how often did you usually eat or drink...'* for nine food items: eight HFSS food items (ready meals, fast food and takeaways, sugary or sweetened drinks, sweets or chocolate, cakes and biscuits, desserts and savoury snacks), and one item for fruit and vegetable intake. Participants were also asked at baseline: '*BEFORE COVID-19, how often did you usually eat or drink...*' for the nine food items. Respondents could answer on a 7-point scale: A few times per day, Once a day, A few times per week, Once a week, A few times per month, Once a month, Less often/never. Apart from for the retrospective 'before COVID-19' question, participants could also answer 'not sure'. To estimate monthly consumption, responses for the food item questions were converted into monthly portion frequencies. Assuming a minimum of four weeks per calendar month, an answer of 'a few times per day' was recorded as 56 portions per month (i.e., 2

portions x 7 days x 4 weeks), 'Once a day' as 28 portions per month, 'Few times per week' as 12 portions per month, 'Once a week' as four portions per month, 'Few times per month' as two portions per month, 'Once a month' as one portion per month, 'Less often/never' as 0.5 portions per month. Participants responding as 'not sure' were excluded.

    - Sugary or sweetened drinks, sweets or chocolate, cakes and biscuits, desserts and savoury snacks were summed to produce a 'HFSS snacks intake' monthly frequency outcome variable. Ready meals, fast food and takeaways monthly frequencies were summed to produce a 'HFSS meals intake' monthly frequency outcome variable.
    - HFSS snacks intake, HFSS meals intake and fruit and vegetable intake at baseline, 3- and 6-months follow-up surveys were combined to create time-varying **HFSS snacks intake** and **HFSS meals intake** and **fruit and vegetable intake** continuous variables.

- **Physical activity (2 levels) 1** (decrease in both or, decrease in either but not an increase in the other), **0** (all other)
    - Physical activity was a binary score comparing strength and aerobic physical activity levels at baseline, 3- and 6-months follow-ups to the retrospectively reported pre-COVID-19 strengthening and aerobic physical activity levels. A reduction in physical activity was defined as a decrease by one or more strength training sessions per week, and/or a decrease by 20 or more minutes per week of aerobic physical activity at each timepoint compared to pre-COVID-19 physical activity levels. A decrease in one, but an increase in the other was treated as 'all other'.
    - **Weekly strength training.** '*BEFORE COVID-19, on average, on HOW MANY DAYS PER* WEEK did you do STRENGTH TRAINING? *'SINCE COVID-19, on average, on HOW MANY DAYS PER WEEK have you done STRENGTH TRAINING?'* 0 days per week, 1 day per week, 2 days per week, 3 days per week, 4 days per week or more. Compared to the before COVID-19 response, a decrease by 1 or more days per week or all other at each timepoint.
    - **Weekly aerobic training**. *'(In the month) BEFORE COVID-19, HOW LONG (in MINUTES) was your average session of moderate or vigorous AEROBIC PHYSICAL ACTIVITY?', 'SINCE COVID-19, HOW LONG (in MINUTES) was your average session of moderate or vigorous AEROBIC PHYSICAL ACTIVITY?'* (15-480 minutes) and '(In the month) BEFORE COVID-19 on average, HOW MANY TIMES PER WEEK did you do at least 15 MINUTES or more of moderate or vigorous AEROBIC PHYSICAL ACTIVITY?', *'SINCE COVID-19 on average, HOW MANY TIMES PER WEEK have you done 15 MINUTES or more of moderate or vigorous AEROBIC PHYSICAL ACTIVITY?'* (0,1,2 to 13 times per week, or 14+ times per week). Compared to the before COVID-19 response, a decrease by 20 or more minutes per week or all other at each timepoint.

- **Alcohol consumption. Binary.** Participants were asked *'How often did you have a drink containing alcohol in the past month?', 'In the past month, how many units of* *alcohol did you drink on a typical day when you were drinking?'.* Scores were multiplied to compute total weekly units of alcohol consumed, and converted into a binary measure of weekly consumption, categorised into **≤14** and **>14** alcohol units.

- **Smoking status. Binary**. At baseline, *'Which statement about tobacco use and cigarette smoking best describes you?'* At follow-up, *'2 MONTHS ago/previously, we asked about your tobacco use. Your situation may have changed since then, or not. Which of the following best applies to you now?'* (2 levels) **Yes** (At baseline: I smoke cigarettes (including hand-rolled) every day, I smoke cigarettes (including hand-rolled), but not every day, I do not smoke cigarettes at all, but I do smoke tobacco of some kind (eg. pipe, cigar or shisha). At follow-up: I smoke cigarettes (including hand-rolled) every day, I smoke cigarettes (including hand-rolled) but not every day, I do not smoke cigarettes at all, but I do smoke tobacco of some kind (e.g. pipe, cigar or shisha)), **No** (At baseline: I have stopped smoking completely in the last year, I stopped smoking completely more than a year ago, I have never smoked any cigarettes. At follow-up: I have stopped smoking or using tobacco completely in the past 2/3 months, I stopped smoking completely more than 2/3 months ago, I have never been a regular smoker (i.e. smoked for a year or more).

**Behavioural, time-invariant**

Time invariant **ΔHFSS snacks intake change score**, **ΔHFSS meals intake change score** and **Δfruit and vegetable intake change score** were computed as described below.

- **ΔHFSS snacks intake change score.** A continuous change score was calculated from the difference between the before-COVID-19 and the since-COVID-19 monthly HFSS snacks frequencies reported at baseline.
- **ΔHFSS meals intake change score.** A continuous change score was calculated from the difference between the before-COVID-19 and the since-COVID-19 monthly HFSS meals frequencies reported at baseline.
- **ΔFruit and vegetable intake change score.** A continuous change score was calculated from the difference between the before-COVID-19 and the since-COVID-19 monthly fruit and vegetable intake frequencies reported at baseline.

**Supplementary tables**

**Table S1.** Weighted baseline participant characteristics for the total, included and excluded samples.
**Table S2.** Means with 95% confidence intervals for weight and BMI at each timepoint using complete cases.
**Table S3.** Mean difference in weight/BMI with 95% confidence intervals between timepoints using complete cases.
**Table S4.** Univariate unadjusted and adjusted models for each predictor variable and change in self-reported weight and BMI.
**Table S5.** Unweighted and weighted proportions of underweight, normal weight, overweight and obesity at baseline (May-June 2020), 3 months (August-September 2020) and 6 months follow-up (November-December 2020) using complete cases.
**Table S6.** Univariate unadjusted and adjusted models for each predictor variable and change in self-reported weight and BMI using complete cases.
**Table S7.** Full GEE model containing all predictor variables and the full GEE model containing all predictor variables and significant predictor*time interactions from univariate models adjusted for time using complete cases.
**Table S8.** Univariate unadjusted and adjusted models for each predictor variable and change in self-reported weight as a binary outcome using complete cases.
**Table S9.** Full GEE model containing all predictor variables and the full GEE model containing all predictor variables and significant predictor*time interactions from univariate models adjusted for time, for a change in self-reported weight as a binary outcome using complete cases.
**Table S10.** Univariate unadjusted and adjusted models for each predictor variable and change in self-reported BMI as a binary outcome using complete cases.
**Table S11.** Full GEE model containing all predictor variables and the full GEE model containing all predictor variables and significant predictor*time interactions from univariate models adjusted for time, for a change in self-reported BMI as a binary outcome using complete cases.

**Table S1.** Weighted baseline participant characteristics for the total, included and excluded samples.

| | **Total Sample** | **Included sample** | **Excluded sample** | |
|---|---|---|---|---|
| | **Weighted %** | **Weighted %** | **Weighted %** | **p** |
| **N** | 2805 | 1631 | 1174 | |
| **Gender** | | | | 0.120 |
| All other | 51.7 | 53.0 | 50.0 | |
| Female | 48.3 | 47.0 | 50.0 | |
| **Ethnicity** | | | - | **<0.001** |
| All other | 11.5 | 7.5 | 17.1 | |
| White | 88.5 | 92.5 | 82.9 | |
| **Mean BMI [SD] (N=2614)** | 26.4 [5.2] | 26.7 [5.1] | 26.0 [5.3] | **0.001** |
| **Mean Age [SD]** | 48.3 [16.7] | 53.1 [14.8] | 41.6 [16.8] | **<0.001** |
| **Occupation and work from home** | | | | **<0.001** |
| Unemployed (including retired persons and students) | 30.9 | 36.2 | 23.5 | |
| Employed working from home | 40.4 | 35.8 | 46.8 | |
| Employed not working from home | 28.7 | 28.0 | 29.6 | |
| **Socioeconomic score** | | | | **<0.001** |
| Income <£50k, unowned Housing and no higher education | 13.0 | 11.5 | 15.2 | |
| 1 of: ≥£50K income, housing ownership/mortgage or higher education | 40.5 | 38.6 | 43.2 | |
| 2 of: ≥£50K income, housing ownership/mortgage or higher education | 34.7 | 37.2 | 31.2 | |
| All of: ≥£50K income, housing ownership/mortgage and higher education | 11.8 | 12.8 | 10.5 | |
| **Living conditions** | | | | **<0.001** |

| | | | | |
|---|---|---|---|---|
| Alone | 18.8% | 19.3% | 18.2% | |
| With children (with or without adults) | 17.3% | 14.8% | 20.7% | |
| With adults only | 63.9% | 65.9% | 61.1% | |
| **Isolation status (N=2763)** | | | | **0.027** |
| Total or some isolation | 75.8% | 77.3% | 73.6% | |
| General or no isolation | 24.2% | 22.7% | 26.4% | |
| **Mean Quality of Life [SD] (1-5) (N=2704)** | 3.2 [0.9] | 3.3 [0.9] | 3.1 [0.9] | **<0.001** |
| **Mean HFSS snacks (portions per month) [SD] (N=2395}** | 61.1 [49.9] | 60.9 [50.7] | 61.5 [48.5] | 0.790 |
| **Mean HFSS meals (portions per month) [SD] (N=2409)** | 7.4 [9.2] | 6.6 [7.4] | 8.8 [11.5] | **<0.001** |
| **Mean Fruit and vegetables (portions per month) [SD] (N=2438)** | 39.4 [19.7] | 40.6 [19.2] | 37.5 [20.2] | **<0.001** |
| **Mean ΔHFSS snacks change score (portions per month) [SD] (N=2395)** | 8.3 [38.8] | 9.8 [39.5] | 5.9 [37.3] | **0.018** |
| **Mean ΔHFSS meals change score (portions per month) [SD] (N=2409)** | -1.4 [8.4] | -1.5 [7.8] | -1.3 [9.4] | 0.649 |
| **Mean ΔFruit and vegetables change score (portions per month) [SD] (N=2438)** | 0.2 [12.6] | 0.1 [11.7] | 0.3 [14.0] | 0.751 |
| **Physical activity (N=2618)** | | | | 0.679 |
| All other | 75.8% | 75.5% | 76.2% | |
| Reduced | 24.2% | 24.5% | 23.8% | |
| **Alcohol consumption (N=2541)** | | | | 0.436 |
| ≤14 weekly units | 80.8% | 81.3% | 80.0% | |
| >14 weekly units | 19.2% | 18.7% | 20.0% | |
| **Smoking status** | | | | **<0.001** |
| Yes | 26.5% | 19.7% | 35.9% | |
| No | 68.9% | 80.3% | 64.1% | |

*SD: Standard Deviation.*

**Table S2.** Means with 95% confidence intervals for weight and BMI at each timepoint using complete cases.

| Complete Cases | | Self-reported weight | | Self-reported BMI | |
|---|---|---|---|---|---|
| | **N** | **Mean [95% CI]** | **SE** | **Mean [95% CI]** | **SE** |
| **May-June 2020** | 1543 | 74.8 [74.0,75.6] | 0.41408 | 26.2 [25.9,26.4] | 0.12693 |
| **August-September 2020** | 1543 | 75.1 [74.3,76.0] | 0.41781 | 26.3 [26.0,26.6] | 0.12905 |
| **November-December 2020** | 1543 | 74.9 [74.0,75.7] | 0.41442 | 26.2 [26.0,26.5] | 0.12820 |

*SE: Standard Error. CI: Confidence Interval.*

**Table S3.** Mean difference in weight/BMI with 95% confidence intervals between timepoints using complete cases.

| Complete Cases | Self-reported weight (kg) | | | | Self-reported BMI (kg/m²) | | |
|---|---|---|---|---|---|---|---|
| | N | Mean difference [95% CI] | SE | p | Mean difference [95% CI] | SE | p |
| **Baseline - 3 mo.** | 1543 | **0.37 [0.14,0.60]** | **0.09514** | **<0.001** | **0.13 [0.06,0.21]** | **0.03319** | **<0.001** |
| **3 mo. - 6 mo.** | 1543 | **-0.28 [-0.48,-0.09]** | **0.08603** | **0.002** | **-0.10 [-0.16,-0.03]** | **0.03000** | **0.002** |
| **Baseline - 6 mo.** | 1543 | 0.08 [-0.14,0.30] | 0.11253 | 0.469 | 0.04 [-0.04,0.11] | 0.03970 | 0.354 |

*SE: Standard Error, CI: Confidence Interval, mo: months.*

**Table S4.** Univariate unadjusted and adjusted models for each predictor variable and change in self-reported weight and BMI.

| | **Change in self-reported weight** | | | | **Change in self-reported BMI** | | | |
|---|---|---|---|---|---|---|---|---|
| | **Unadjusted models** | | **Adjusted models w/interaction** | | **Unadjusted models** | | **Adjusted models w/interaction** | |
| **Predictor** | **W χ2** | **p** | **W χ2** | **p** | **W χ2** | **p** | **W χ2** | **p** |
| Gender | 2.030 | 0.154 | 1.883 | 0.170 | 2.591 | 0.107 | 2.469 | 0.116 |
| Time | | | **10.243** | **0.001** | | | **10.632** | **0.001** |
| Time*Gender | | | 1.669 | 0.196 | | | 1.176 | 0.278 |
| Ethnicity | 0.069 | 0.792 | 0.034 | 0.853 | 0.074 | 0.785 | 0.038 | 0.845 |
| Time | | | **5.811** | **0.016** | | | **5.890** | **0.015** |
| Time*Ethnicity | | | 0.779 | 0.377 | | | 0.910 | 0.340 |
| Baseline BMI | **11.759** | **0.001** | **10.529** | **0.001** | **11.353** | **0.001** | **10.187** | **0.001** |
| Time | | | **7.856** | **0.005** | | | **7.563** | **0.006** |
| Time*Baseline BMI | | | **11.001** | **0.001** | | | **10.596** | **0.001** |
| Age | 0.231 | 0.631 | 0.146 | 0.702 | 0.165 | 0.684 | 0.094 | 0.760 |
| Time | | | 2.145 | 0.143 | | | 2.237 | 0.135 |
| Time*Age | | | 0.459 | 0.498 | | | 0.491 | 0.483 |
| Occupation and work from home | 0.313 | 0.855 | 0.280 | 0.869 | 0.198 | 0.906 | 0.179 | 0.914 |
| Time | | | **8.504** | **0.004** | | | **8.365** | **0.004** |
| Time*Occupation and work from home | | | 0.488 | 0.783 | | | 0.279 | 0.870 |

| | | | | | | | | |
|---|---|---|---|---|---|---|---|---|
| Socioeconomic score | 2.514 | 0.473 | 2.338 | 0.505 | 2.891 | 0.409 | 2.696 | 0.441 |
| Time | | | 3.703 | 0.054 | | | 3.515 | 0.061 |
| Time*Socioeconomic score | | | 2.073 | 0.557 | | | 2.394 | 0.495 |
| Living arrangements | 0.344 | 0.842 | 0.389 | 0.823 | 0.430 | 0.806 | 0.470 | 0.791 |
| Time | | | **4.445** | **0.035** | | | **4.985** | **0.026** |
| Time*Living arrangements | | | 0.870 | 0.647 | | | 0.557 | 0.757 |
| Isolation status | 0.680 | 0.410 | 0.448 | 0.503 | 0.634 | 0.426 | 0.533 | 0.465 |
| Time | | | **12.203** | **<0.001** | | | **12.975** | **<0.001** |
| Time*Isolation status | | | 0.978 | 0.323 | | | 1.379 | 0.240 |
| Quality of life | 0.174 | 0.676 | 0.038 | 0.846 | 0.219 | 0.640 | 0.023 | 0.878 |
| Time | | | 0.259 | 0.611 | | | 0.411 | 0.522 |
| Time*Quality of life | | | 0.008 | 0.928 | | | 0.003 | 0.955 |
| HFSS snacks intake | **15.593** | **<0.001** | **13.775** | **<0.001** | **15.772** | **<0.001** | **14.328** | **<0.001** |
| Time | | | **29.291** | **<0.001** | | | **28.201** | **<0.001** |
| Time*HFSS snacks intake | | | **13.221** | **<0.001** | | | **12.206** | **<0.001** |
| HFSS meals intake | 2.002 | 0.157 | 1.851 | 0.174 | 1.600 | 0.206 | 1.321 | 0.250 |
| Time | | | **19.85** | **<0.001** | | | **21.041** | **<0.001** |
| Time*HFSS meals intake | | | **4.257** | **0.039** | | | **4.839** | **0.028** |

| | | | | | | | | |
|---|---|---|---|---|---|---|---|---|
| Fruit and vegetables intake | **5.397** | **0.020** | **5.456** | **0.020** | **5.315** | **0.021** | **5.425** | **0.020** |
| Time | | | 0.792 | 0.374 | | | 0.855 | 0.355 |
| Time*Fruit and vegetables intake | | | **4.475** | **0.034** | | | **4.776** | **0.029** |
| ∆HFSS snacks change score | **4.296** | **0.038** | **4.532** | **0.033** | **4.599** | **0.032** | **4.881** | **0.027** |
| Time | | | **7.368** | **0.007** | | | **7.003** | **0.008** |
| Time*∆HFSS snacks change score | | | 0.185 | 0.667 | | | 0.199 | 0.655 |
| ∆HFSS meals change score | 0.008 | 0.929 | 0.004 | 0.950 | 0.003 | 0.954 | 0.001 | 0.979 |
| Time | | | **10.258** | **0.001** | | | **10.276** | **0.001** |
| Time*∆HFSS meals change score | | | 0.018 | 0.894 | | | 0.024 | 0.878 |
| ∆Fruit and vegetables change score | 1.037 | 0.309 | 1.080 | 0.299 | 1.303 | 0.254 | 1.356 | 0.244 |
| Time | | | **9.285** | **0.002** | | | **9.084** | **0.003** |
| Time*∆Fruit and vegetables change score | | | 0.025 | 0.875 | | | 0.038 | 0.845 |
| Physical activity | 0.002 | 0.963 | <0.001 | 0.999 | 0.032 | 0.859 | 0.051 | 0.821 |
| Time | | | **7.562** | **0.006** | | | **7.110** | **0.008** |
| Time*Physical activity | | | 0.236 | 0.627 | | | 0.341 | 0.559 |
| Alcohol consumption | 2.946 | 0.086 | 2.373 | 0.123 | 2.289 | 0.130 | 1.778 | 0.182 |
| Time | | | 1.514 | 0.219 | | | 1.751 | 0.186 |
| Time*Alcohol consumption | | | **9.351** | **0.002** | | | **8.974** | **0.003** |
| Smoking status | 0.092 | 0.762 | 0.072 | 0.789 | 0.092 | 0.762 | 0.071 | 0.790 |

| | | | | |
|---|---|---|---|---|
| Time | 2.854 | 0.091 | 2.906 | 0.088 |
| Time*Smoking status | 0.077 | 0.782 | 0.079 | 0.778 |

*Missingness with each model depends on if participants responded to survey questions relating to the predictor variable. W $\chi2$: Wald Chi-square.*

**Table S5.** Unweighted and weighted proportions of underweight, normal weight, overweight and obesity at baseline (May-June 2020), 3 months (August-September 2020) and 6 months follow-up (November-December 2020) using complete cases.

| | Unweighted | | | Weighted | | |
|---|---|---|---|---|---|---|
| | **Apr-Jun** | **Jul-Sep** | **Nov-Dec** | **Apr-Jun** | **Jul-Sep** | **Nov-Dec** |
| **N** | 1543 | 1543 | 1543 | 1333 | 1392 | 1358 |
| **BMI Category (%)** | | | | | | |
| Underweight | 2.0 | 2.0 | 2.1 | 2.6 | 2.9 | 2.6 |
| Normal weight | 45.0 | 44.1 | 44.9 | 38.3 | 38.8 | 39.9 |
| Overweight | 32.8 | 32.9 | 32.8 | 37.0 | 35.7 | 35.5 |
| Obesity | 20.2 | 21.0 | 20.2 | 22.1 | 22.7 | 22.0 |

**Table S6.** Univariate unadjusted and adjusted models for each predictor variable and change in self-reported weight and BMI using complete cases.

| | Change in self-reported weight | | | | Change in self-reported BMI | | | |
|---|---|---|---|---|---|---|---|---|
| | Unadjusted models | | Adjusted models w/interaction | | Unadjusted models | | Adjusted models w/interaction | |
| **Predictor** | **W χ2** | **p** | **W χ2** | **p** | **W χ2** | **p** | **W χ2** | **p** |
| Gender | 2.761 | 0.097 | 2.761 | 0.097 | 3.287 | 0.070 | 3.287 | 0.070 |
| Time | | | **11.000** | **0.001** | | | **11.232** | **0.001** |
| Time*Gender | | | 2.017 | 0.156 | | | 1.429 | 0.232 |
| Ethnicity | 0.001 | 0.978 | 0.001 | 0.978 | 0.012 | 0.912 | 0.012 | 0.912 |
| Time | | | **5.915** | **0.015** | | | **6.159** | **0.013** |
| Time*Ethnicity | | | 0.396 | 0.529 | | | 0.669 | 0.413 |
| Baseline BMI | **9.913** | **0.002** | **9.913** | **0.002** | **9.680** | **0.002** | **9.680** | **0.002** |
| Time | | | **8.070** | **0.005** | | | **7.835** | **0.005** |
| Time*Baseline BMI | | | **11.286** | **0.001** | | | **10.918** | **0.001** |
| Age | 0.354 | 0.552 | 0.354 | 0.552 | 0.344 | 0.558 | 0.344 | 0.558 |
| Time | | | 2.390 | 0.122 | | | 2.537 | 0.111 |
| Time*Age | | | 0.574 | 0.449 | | | 0.642 | 0.423 |
| Occupation and work from home | 0.888 | 0.642 | 0.888 | 0.642 | 0.738 | 0.691 | 0.738 | 0.691 |
| Time | | | **8.411** | **0.004** | | | **8.155** | **0.004** |

| | | | | | | | | |
|---|---|---|---|---|---|---|---|---|
| Time*Occupation and work from home | | | 0.804 | 0.669 | | | 0.552 | 0.759 |
| Socioeconomic score | 1.087 | 0.780 | 1.087 | 0.780 | 1.395 | 0.707 | 1.395 | 0.707 |
| Time | | | 3.878 | 0.049 | | | 3.403 | 0.065 |
| Time*Socioeconomic score | | | 1.553 | 0.670 | | | 1.905 | 0.592 |
| Living arrangements | 0.510 | 0.775 | 0.510 | 0.775 | 0.730 | 0.694 | 0.730 | 0.694 |
| Time | | | **5.219** | **0.022** | | | **5.691** | **0.017** |
| Time*Living arrangements | | | 0.803 | 0.669 | | | 0.526 | 0.769 |
| Isolation status | 1.058 | 0.304 | 0.250 | 0.617 | 0.939 | 0.332 | 0.327 | 0.567 |
| Time | | | **12.113** | **0.001** | | | **12.802** | **<0.001** |
| Time*Isolation status | | | 1.233 | 0.267 | | | 1.633 | 0.201 |
| Quality of life | 0.036 | 0.850 | 0.216 | 0.642 | 0.053 | 0.819 | 0.171 | 0.679 |
| Time | | | 0.138 | 0.710 | | | 0.248 | 0.618 |
| Time*Quality of life | | | 0.058 | 0.809 | | | 0.009 | 0.925 |
| HFSS snacks intake | **13.217** | **<0.001** | **13.025** | **<0.001** | **12.956** | **<0.001** | **13.064** | **<0.001** |
| Time | | | **29.276** | **<0.001** | | | **28.001** | **<0.001** |
| Time*HFSS snacks intake | | | **13.198** | **<0.001** | | | **12.030** | **0.001** |
| HFSS meals intake | 2.522 | 0.112 | 2.878 | 0.090 | 1.732 | 0.188 | 1.927 | 0.165 |
| Time | | | **21.490** | **<0.001** | | | **22.116** | **<0.001** |
| Time*HFSS meals intake | | | **4.842** | **0.028** | | | **5.124** | **0.024** |

| | | | | | | | | |
|---|---|---|---|---|---|---|---|---|
| Fruit and vegetables intake | **6.377** | **0.012** | **6.922** | **0.009** | **6.206** | **0.013** | **6.772** | **0.009** |
| Time | | | 1.136 | 0.287 | | | 1.207 | 0.272 |
| Time*Fruit and vegetables intake | | | **5.352** | **0.021** | | | **5.671** | **0.017** |
| ∆HFSS snacks change score | 2.058 | 0.151 | 2.058 | 0.151 | 2.177 | 0.140 | 2.177 | 0.140 |
| Time | | | **7.896** | **0.005** | | | **7.424** | **0.006** |
| Time*∆HFSS snacks change score | | | 0.436 | 0.509 | | | 0.452 | 0.502 |
| ∆HFSS meals change score | 0.544 | 0.461 | 0.544 | 0.461 | 0.389 | 0.533 | 0.389 | 0.533 |
| Time | | | **12.149** | **<0.001** | | | **11.977** | **0.001** |
| Time*∆HFSS meals change score | | | 0.159 | 0.691 | | | 0.139 | 0.709 |
| ∆Fruit and vegetables change score | 0.122 | 0.727 | 0.122 | 0.727 | 0.199 | 0.655 | 0.199 | 0.655 |
| Time | | | **10.692** | **0.001** | | | **10.345** | **0.001** |
| Time*∆Fruit and vegetables change score | | | 0.031 | 0.861 | | | 0.019 | 0.890 |
| Physical activity | 0.150 | 0.903 | **0.010** | 0.920 | 0.015 | 0.903 | 0.020 | 0.887 |
| Time | | | **7.685** | **0.006** | | | **7.215** | **0.007** |
| Time*Physical activity | | | 0.30 | 0.584 | | | 0.398 | 0.528 |
| Alcohol consumption | 2.122 | 0.145 | 1.934 | 0.164 | 1.674 | 0.196 | 1.503 | 0.220 |
| Time | | | 2.136 | 0.144 | | | 2.357 | 0.125 |
| Time*Alcohol consumption | | | **8.811** | **0.003** | | | **8.527** | **0.003** |

| | | | | | | | | |
|---|---|---|---|---|---|---|---|---|
| Smoking status | 0.066 | 0.797 | 0.052 | 0.820 | 0.048 | 0.826 | 0.036 | 0.850 |
| Time | | | 2.910 | 0.088 | | | 2.872 | 0.090 |
| Time*Smoking status | | | 0.079 | 0.778 | | | 0.091 | 0.762 |

*Missingness with each model depends on if participants responded to survey questions relating to the predictor variable. W χ2: Wald Chi-square.*

**Table S7.** Full GEE model containing all predictor variables and the full GEE model containing all predictor variables and significant predictor*time interactions from univariate models adjusted for time using complete cases.

| Complete cases | **Change in self-reported weight QIC = 43324.151** | | | | **Change in self-reported BMI QIC = 5392.741** | | | |
|---|---|---|---|---|---|---|---|---|
| All predictors (N=1416) | **W χ2** | **p** | **B [95% CI]** | **SE** | **W χ2** | **p** | **B [95% CI]** | **SE** |
| Gender | 3.073 | 0.080 | | | 3.564 | 0.059 | | |
| All other | | | Reference | | | | Reference | |
| Female | | | 0.418 [-0.049,0.886] | 0.2387 | | | 0.147 [-0.006,0.300] | 0.0780 |
| Ethnicity | 0.072 | 0.788 | | | 0.056 | 0.812 | | |
| All other | | | Reference | | | | Reference | |
| White | | | -0.088 [-0.728,0.553] | 0.3268 | | | -0.023 [-0.268,0.215] | 0.1252 |
| Baseline BMI | **9.865** | **0.002** | **-0.091 [-0.148,-0.034]** | **0.0291** | **9.823** | **0.002** | **-0.032 [-0.052,-0.012]** | **0.0103** |
| Age | 1.814 | 0.178 | 0.011 [-0.005,0.026] | 0.0080 | 2.091 | 0.148 | 0.004 [-0.001,0.009] | 0.0028 |
| Occupation and work from home | 0.058 | 0.971 | | | 0.011 | 0.995 | | |
| Unemployed | | | Reference | | | | Reference | |
| Employed working from home | | | -0.068 [-0.619,0.484] | 0.2814 | | | -0.010 [-0.206,0.185] | 0.0998 |
| Employed not working from home | | | -0.049 [-0.722,0.624] | 0.3434 | | | -0.008 [-0.242,0.227] | 0.1197 |
| Socioeconomic score | 1.448 | 0.694 | | | 1.650 | 0.648 | | |
| Income <£50k, unowned Housing and no higher education | | | Reference | | | | Reference | |
| 1 of: ≥£50K income, housing ownership/mortgage or higher education | | | 0.523 [-0.815,1.860] | 0.6826 | | | 0.111 [-0.349,0.570] | 0.2346 |
| 2 of: ≥£50K income, housing ownership/mortgage or higher education | | | 0.282 [-0.990,1.554] | 0.6490 | | | 0.015 [-0.420,0.449] | 0.2218 |
| All of: ≥£50K income, housing ownership/mortgage and higher education | | | 0.152 [-1.148,1.453] | 0.6635 | | | -0.033 [-0.477,0.411] | 0.2267 |
| Living conditions | 0.100 | 0.951 | | | 0.135 | 0.935 | | |
| Alone | | | Reference | | | | Reference | |
| With children (with or without adults) | | | 0.097 [-0.532,0.725] | 0.3206 | | | 0.036 [-0.185,0.256] | 0.1124 |
| With adults only | | | 0.072 [-0.440,0.584] | 0.2611 | | | 0.033 [-0.148,0.214] | 0.0923 |
| Isolation status | 0.339 | 0.561 | | | 0.373 | 0.541 | | |
| Total or some isolation | | | Reference | | | | Reference | |

| | | | | | | | | |
|---|---|---|---|---|---|---|---|---|
| General or no isolation | | | -0.092 [-0.403,0.218] | 0.1584 | | | -0.034 [-0.143,0.075] | 0.0558 |
| Quality of Life | <0.001 | 0.996 | <0.001 [-0.220,0.219] | 0.1121 | 0.015 | 0.904 | 0.005 [-0.075,0.085] | 0.0406 |
| HFSS snacks | **11.656** | **0.001** | **0.009 [0.004,0.014]** | **0.0027** | **11.929** | **0.001** | **0.003 [0.001,0.005]** | **0.0010** |
| HFSS meals | 3.677 | 0.055 | 0.021 [<0.001,0.043] | 0.0110 | 2.777 | 0.096 | 0.006 [-0.001,0.014] | 0.0039 |
| Fruit and vegetables | **5.215** | **0.022** | **-0.011 [-0.021,-0.002]** | **0.0050** | **5.043** | **0.025** | **-0.004 [-0.007,<0.001]** | **0.0018** |
| ∆HFSS snacks change score | 2.054 | 0.152 | 0.008 [-0.003,0.019] | 0.0057 | 2.228 | 0.136 | 0.003 [-0.001,0.006] | 0.0018 |
| ∆HFSS meals change score | 0.366 | 0.545 | -0.010 [-0.044,0.023] | 0.0172 | 0.285 | 0.593 | -0.003 [-0.015,0.008] | 0.0058 |
| ∆Fruit and vegetables change score | 0.062 | 0.804 | -0.002 [-0.020,0.016] | 0.0091 | 0.090 | 0.764 | -0.001 [-0.007,0.005] | 0.0031 |
| Physical activity | 0.031 | 0.861 | | | 0.007 | 0.932 | | |
| All other | | | Reference | | | | Reference | |
| Reduced | | | 0.028 [-0.283,0.339] | 0.1586 | | | -0.005 [-0.115,0.105] | 0.0560 |
| Alcohol consumption | 3.707 | 0.054 | | | 3.330 | 0.068 | | |
| ≤14 weekly units | | | Reference | | | | Reference | |
| >14 weekly units | | | 0.390 [-0.007,0.787] | 0.2024 | | | 0.121 [-0.009,0.252] | 0.0665 |
| Smoking status | 0.045 | 0.833 | | | 0.018 | 0.893 | | |
| Yes | | | Reference | | | | Reference | |
| No | | | 0.064 [-0.528,0.656] | 0.3020 | | | 0.014 [-0.196,.225] | 0.1072 |

| | **Change in self-reported weight QIC = 43090.196** | | **Change in self-reported BMI QIC = 5366.550** | |
|---|---|---|---|---|
| All predictors + significant time interactions (N=1416) | **W χ2** | **p** | **W χ2** | **p** |
| Time*Baseline BMI | **13.202** | **<0.001** | **12.463** | **<0.001** |
| Time*HFSS snacks | **16.997** | **<0.001** | **15.634** | **<0.001** |
| Time*Alcohol consumption | **12.908** | **<0.001** | **12.66** | **0.001** |

*Models also included Time as a covariate. For 'All predictors + significant time interactions', Type III tests for the predictor*time interactions are shown only. There were no material changes in significance of main effects with the addition of the time interactions, except for alcohol consumption (p=0.046) and HFSS meals intake (p=0.049) which became significant for a change in weight. QIC is a relative, 'lower is better' measure of goodness of fit. W χ2: Wald Chi-square, $B$: Beta parameter, SE: Standard Error, CI: Confidence Interval.*

**Table S8.** Univariate unadjusted and adjusted models for each predictor variable and change in self-reported weight as a binary outcome using complete cases.

| Binary Weight | Increase weight vs all other | | | | Decrease weight vs all other | | | |
|---|---|---|---|---|---|---|---|---|
| | **Unadjusted models** | | **Adjusted models w/interaction** | | **Unadjusted models** | | **Adjusted models w/interaction** | |
| **Predictor** | **W χ2** | **p** | **W χ2** | **p** | **W χ2** | **p** | **W χ2** | **p** |
| Gender | 1.064 | 0.302 | 1.011 | 0.315 | 3.034 | 0.082 | 2.925 | 0.087 |
| Time | | | 1.512 | 0.219 | | | **30.785** | **<0.001** |
| Time*Gender | | | 0.925 | 0.336 | | | 0.019 | 0.889 |
| Ethnicity | 0.002 | 0.969 | 0.002 | 0.968 | 0.686 | 0.407 | ** | ** |
| Time | | | <0.001 | 0.992 | | | ** | ** |
| Time*Ethnicity | | | 0.420 | 0.517 | | | ** | ** |
| Baseline BMI | 2.290 | 0.130 | 2.322 | 0.128 | **13.351** | **<0.001** | **12.163** | **<0.001** |
| Time | | | 0.666 | 0.414 | | | 28.750 | **<0.001** |
| Time*Baseline BMI | | | 0.222 | 0.638 | | | 0.063 | 0.802 |
| Age | **5.106** | **0.024** | **5.146** | **0.023** | 1.282 | 0.257 | 1.303 | 0.254 |
| Time | | | 0.113 | 0.736 | | | 1.880 | 0.170 |
| Time*Age | | | 0.032 | 0.859 | | | 0.076 | 0.783 |
| Occupation and work from home | 0.912 | 0.634 | 0.834 | 0.659 | 0.731 | 0.694 | 1.402 | 0.496 |
| Time | | | **4.035** | **0.045** | | | **25.886** | **<0.001** |
| Time*Occupation and work from home | | | 1.506 | 0.471 | | | 4.449 | 0.108 |

| | | | | | | | | |
|---|---|---|---|---|---|---|---|---|
| Socioeconomic score | **14.341** | **0.002** | **13.948** | **0.003** | 6.622 | 0.085 | ** | ** |
| Time | | | 2.528 | 0.112 | | | ** | ** |
| Time*Socioeconomic score | | | 1.847 | 0.605 | | | ** | ** |
| Living arrangements | 1.140 | 0.565 | 1.105 | 0.576 | 1.878 | 0.391 | 1.462 | 0.481 |
| Time | | | **2.948** | **0.086** | | | **25.406** | **<0.001** |
| Time*Living arrangements | | | 0.883 | 0.643 | | | 0.367 | 0.832 |
| Isolation status | 0.064 | 0.800 | 0.076 | 0.783 | **9.201** | **0.002** | 0.427 | 0.513 |
| Time | | | 1.846 | 0.174 | | | **20.967** | **<0.001** |
| Time*Isolation status | | | 1.288 | 0.256 | | | 0.308 | 0.579 |
| Quality of life | 2.156 | 0.142 | 1.360 | 0.244 | **9.909** | **0.002** | **6.578** | **0.010** |
| Time | | | 0.138 | 0.711 | | | 0.610 | 0.435 |
| Time*Quality of life | | | 0.522 | 0.470 | | | **4.123** | **0.042** |
| HFSS snacks | 3.350 | 0.067 | 1.000 | 0.317 | **4.307** | **0.038** | 3.636 | 0.057 |
| Time | | | 0.470 | 0.493 | | | **34.552** | **<0.001** |
| Time*HFSS snacks | | | **3.996** | **0.046** | | | 0.911 | 0.340 |
| HFSS meals | 1.814 | 0.178 | 0.488 | 0.485 | 0.001 | 0.980 | 0.009 | 0.924 |
| Time | | | 0.003 | 0.959 | | | **29.907** | **<0.001** |
| Time*HFSS meals | | | 2.790 | 0.095 | | | 3.059 | 0.080 |

| | | | | | | | | |
|---|---|---|---|---|---|---|---|---|
| Fruit and vegetables | **13.939** | **<0.001** | **12.561** | **<0.001** | 0.989 | 0.320 | 0.813 | 0.367 |
| Time | | | **1.458** | **0.227** | | | 17.885 | <0.001 |
| Time*Fruit and vegetables | | | 0.566 | 0.452 | | | 2.322 | 0.128 |
| ΔHFSS snacks change score | 2.848 | 0.091 | 2.832 | 0.092 | 0.415 | 0.519 | 0.704 | 0.401 |
| Time | | | 2.119 | 0.145 | | | 31.601 | <0.001 |
| Time*ΔHFSS snacks change score | | | 0.006 | 0.937 | | | 1.280 | 0.258 |
| ΔHFSS meals change score | 0.154 | 0.695 | 0.138 | 0.710 | 2.428 | 0.119 | 1.866 | 0.172 |
| Time | | | 1.768 | 0.184 | | | **35.365** | **<0.001** |
| Time*ΔHFSS meals change score | | | 0.147 | 0.701 | | | 1.633 | 0.201 |
| ΔFruit and vegetables change score | 0.184 | 0.668 | 0.177 | 0.674 | 0.011 | 0.916 | 0.027 | 0.869 |
| Time | | | 2.480 | 0.115 | | | **33.273** | **<0.001** |
| Time*ΔFruit and vegetables change score | | | 0.006 | 0.938 | | | 0.341 | 0.559 |
| Physical activity | 0.850 | 0.357 | 0.763 | 0.382 | 0.321 | 0.571 | 0.019 | 0.890 |
| Time | | | 1.429 | 0.232 | | | **24.086** | **<0.001** |
| Time*Physical activity | | | 0.978 | 0.323 | | | 2.438 | 0.118 |
| Alcohol consumption | 0.027 | 0.870 | 0.005 | 0.941 | **5.755** | **0.016** | **4.314** | **0.038** |
| Time | | | 2.934 | 0.087 | | | **12.509** | **<0.001** |
| Time*Alcohol consumption | | | 0.838 | 0.360 | | | **4.854** | **0.028** |
| Smoking status | 2.974 | 0.085 | 2.774 | 0.096 | **8.076** | **0.004** | **7.192** | **0.007** |

| | | | | | | |
|---|---|---|---|---|---|---|
| Time | 2.901 | 0.089 | | | **13.501** | **<0.001** |
| Time*Smoking status | 0.227 | 0.634 | | | 1.705 | 0.192 |

***Model did not compute due to numerical errors. In the 'Decrease weight vs all other' model, baseline BMI, HFSS snacks intake, fruit and vegetable intake, and HFSS snacks change score continuous variables were converted to categorical variables due to violating the linearity of logit assumption. Missingness with each model depends on if participants responded to survey questions relating to the predictor variable. W χ2: Wald Chi-square.*

**Table S9.** Full GEE model containing all predictor variables and the full GEE model containing all predictor variables and significant predictor*time interactions from univariate models adjusted for time, for a change in self-reported weight as a binary outcome using complete cases.

| **Binary weight outcome variable** | | **Increase weight vs all other** | | | **Decrease weight vs all other** | | |
|---|---|---|---|---|---|---|---|
| | | | **Type III Test** | | | **Type III Test** | |
| **Model** | **Predictor** | **QIC** | **W χ2** | **p** | **QIC** | **W χ2** | **p** |
| All predictors (N=1416) | | 1806.499 | | | 1416.847 | | |
| | Gender | | 1.037 | 0.309 | | **7.545** | **0.006** |
| | Ethnicity | | 0.008 | 0.928 | | 0.766 | 0.381 |
| | Baseline BMI | | 1.922 | 0.166 | | **13.646** | **<0.001** |
| | Age | | **6.570** | **0.010** | | 2.856 | 0.091 |
| | Occupation and work from home | | 3.829 | 0.147 | | 0.848 | 0.654 |
| | Socioeconomic score | | 4.761 | 0.190 | | 3.461 | 0.326 |
| | Living conditions | | 0.876 | 0.645 | | 1.402 | 0.496 |
| | Isolation status | | 0.002 | 0.963 | | 0.837 | 0.360 |
| | Quality of Life | | 0.069 | 0.792 | | 1.361 | 0.243 |
| | HFSS snacks intake | | 1.519 | 0.218 | | **5.345** | **0.021** |
| | HFSS meals intake | | 0.019 | 0.890 | | 1.040 | 0.308 |
| | Fruit and vegetables intake | | **9.368** | **0.002** | | 3.000 | 0.083 |
| | ΔHFSS snacks change score | | 3.585 | 0.058 | | 0.105 | 0.746 |
| | ΔHFSS meals change score | | 0.333 | 0.564 | | 2.047 | 0.152 |
| | ΔFruit and vegetables change score | | 0.243 | 0.622 | | 0.012 | 0.912 |
| | Physical activity | | 0.674 | 0.412 | | 0.016 | 0.899 |
| | Alcohol consumption | | 0.022 | 0.883 | | **11.158** | **0.001** |
| | Smoking status | | 0.026 | 0.873 | | 2.306 | 0.129 |

| | | | | | | | |
|---|---|---|---|---|---|---|---|
| All predictors and significant time interactions (N=1416) | | 1803.355 | | | 1416.522 | | |
| | Time*HFSS snacks intake | | 3.775 | 0.052 | | | |
| | Time*Alcohol consumption | | | | | **5.078** | **0.024** |

*Models also included Time as a covariate. Type III tests are shown only. There were no material changes in significance of main effects. In the ‘Decrease weight vs all other’ model, baseline BMI, HFSS snacks intake, fruit and vegetable intake, and HFSS snacks change score continuous variables were converted to categorical variables due to violating the linearity of logit assumption. QIC is a relative, ‘lower is better’ measure of goodness of fit. W χ2: Wald Chi-square.*

**Table S10.** Univariate unadjusted and adjusted models for each predictor variable and change in self-reported BMI as a binary outcome using complete cases.

| Binary BMI outcome variable | **Increase BMI vs all other (<25 BMI)** | | | | **Decrease BMI vs all other (≥25 BMI)** | | | |
|---|---|---|---|---|---|---|---|---|
| | **Unadjusted models** | | **Adjusted models w/interaction** | | **Unadjusted models** | | **Adjusted models w/interaction** | |
| **Predictor** | **W χ2** | **p** | **W χ2** | **p** | **W χ2** | **p** | **W χ2** | **p** |
| Gender | 0.005 | 0.945 | 0.013 | 0.909 | 0.387 | 0.534 | 0.284 | 0.594 |
| Time | | | **5.261** | **0.022** | | | **23.65** | **<0.001** |
| Time*Gender | | | 0.675 | 0.411 | | | 0.150 | 0.699 |
| Ethnicity | 0.110 | 0.741 | 0.115 | 0.735 | 2.098 | 0.147 | ** | ** |
| Time | | | 0.585 | 0.444 | | | ** | ** |
| Time*Ethnicity | | | 0.014 | 0.906 | | | ** | ** |
| Baseline BMI | **12.444** | **<0.001** | **12.274** | **<0.001** | 1.160 | 0.281 | 0.382 | 0.537 |
| Time | | | 2.712 | 0.100 | | | 1.067 | 0.302 |
| Time*Baseline BMI | | | 0.061 | 0.804 | | | 3.070 | 0.080 |
| Age | 0.507 | 0.476 | 0.294 | 0.588 | 2.094 | 0.148 | 2.449 | 0.118 |
| Time | | | 0.639 | 0.424 | | | 0.032 | 0.858 |
| Time*Age | | | 1.615 | 0.204 | | | 1.233 | 0.267 |
| Occupation and work from home | 1.250 | 0.535 | 1.119 | 0.572 | 2.551 | 0.279 | 3.413 | 0.181 |
| Time | | | 3.553 | 0.059 | | | **21.051** | **<0.001** |

| | | | | | | | | |
|---|---|---|---|---|---|---|---|---|
| Time*Occupation and work from home | | | 1.103 | 0.576 | | | 4.483 | 0.106 |
| Socioeconomic score | **8.415** | **0.038** | 7.772 | 0.051 | 3.773 | 0.287 | 4.420 | 0.220 |
| Time | | | **3.905** | **0.048** | | | **13.021** | **<0.001** |
| Time*Socioeconomic score | | | 2.811 | 0.422 | | | 2.022 | 0.568 |
| Living arrangements | 1.504 | 0.471 | 1.376 | 0.502 | 1.221 | 0.543 | 0.546 | 0.761 |
| Time | | | **4.719** | **0.030** | | | **23.423** | **<0.001** |
| Time*Living arrangements | | | 0.868 | 0.648 | | | 3.006 | 0.222 |
| Isolation status | 2.858 | 0.091 | 0.662 | 0.416 | 2.764 | 0.096 | 0.553 | 0.457 |
| Time | | | 1.988 | 0.159 | | | **18.484** | **<0.001** |
| Time*Isolation status | | | 0.017 | 0.898 | | | 1.885 | 0.170 |
| Quality of life | 0.162 | 0.687 | 0.001 | 0.980 | **5.064** | **0.024** | 2.430 | 0.119 |
| Time | | | 2.208 | 0.137 | | | 0.290 | 0.590 |
| Time*Quality of life | | | 3.74 | 0.053 | | | 0.264 | 0.607 |
| HFSS snacks intake | 1.213 | 0.271 | 0.533 | 0.466 | **5.843** | **0.016** | **6.411** | **0.011** |
| Time | | | 0.493 | 0.483 | | | **15.768** | **<0.001** |
| Time*HFSS snacks intake | | | 0.416 | 0.519 | | | 0.784 | 0.376 |
| HFSS meals intake | 1.095 | 0.295 | 1.281 | 0.258 | 1.512 | 0.219 | 1.204 | 0.273 |
| Time | | | 3.188 | 0.074 | | | **20.456** | **<0.001** |
| Time*HFSS meals intake | | | 0.136 | 0.713 | | | 1.537 | 0.215 |

| | | | | | | | | |
|---|---|---|---|---|---|---|---|---|
| Fruit and vegetables intake | **8.678** | **0.003** | **7.481** | **0.006** | **5.980** | **0.014** | **6.847** | **0.009** |
| Time | | | 0.628 | 0.428 | | | 2.681 | 0.102 |
| Time*Fruit and vegetables intake | | | 0.070 | 0.791 | | | 0.060 | 0.807 |
| ∆HFSS snacks change score | 0.751 | 0.386 | 0.824 | 0.364 | 0.515 | 0.473 | 0.047 | 0.829 |
| Time | | | **5.343** | **0.021** | | | **17.772** | **<0.001** |
| Time*∆HFSS snacks change score | | | 1.898 | 0.168 | | | 2.031 | 0.154 |
| ∆HFSS meals change score | 2.814 | 0.093 | 2.514 | 0.113 | 1.079 | 0.299 | 0.726 | 0.394 |
| Time | | | 3.232 | 0.072 | | | **26.727** | **<0.001** |
| Time*∆HFSS meals change score | | | 0.344 | 0.558 | | | 2.756 | 0.097 |
| ∆Fruit and vegetables change score | 0.052 | 0.820 | 0.079 | 0.778 | 0.054 | 0.816 | 0.065 | 0.799 |
| Time | | | **4.413** | **0.036** | | | **25.119** | **<0.001** |
| Time*∆Fruit and vegetables change score | | | 0.095 | 0.758 | | | 0.041 | 0.840 |
| Physical activity | 0.623 | 0.430 | 0.477 | 0.490 | 0.204 | 0.652 | 0.021 | 0.884 |
| Time | | | 1.716 | 0.190 | | | **15.761** | **<0.001** |
| Time*Physical activity | | | 1.324 | 0.250 | | | 2.465 | 0.116 |
| Alcohol consumption | 0.222 | 0.638 | 0.155 | 0.694 | **6.014** | **0.014** | **4.769** | **0.029** |
| Time | | | 3.336 | 0.068 | | | **8.708** | **0.003** |
| Time*Alcohol consumption | | | 0.475 | 0.491 | | | 2.549 | 0.110 |

| | | | | | | | | |
|---|---|---|---|---|---|---|---|---|
| Smoking status | 0.958 | 0.328 | 0.682 | 0.409 | **5.260** | **0.022** | **4.397** | **0.036** |
| Time | | | **4.700** | **0.030** | | | **9.806** | **0.002** |
| Time*Smoking status | | | 0.766 | 0.381 | | | 1.886 | 0.170 |

***Model did not compute due to numerical errors. In the ‘Increase BMI vs all other’ GEE model, the baseline BMI continuous variable was converted to a categorical variable, due to violating the linearity of logit assumption. Missingness with each model depends on if participants responded to survey questions relating to the predictor variable. W χ2: Wald Chi-square.*

**Table S11.** Full GEE model containing all predictor variables and the full GEE model containing all predictor variables and significant predictor*time interactions from univariate models adjusted for time, for a change in self-reported BMI as a binary outcome using complete cases.

| **Binary BMI outcome variable** | | **Increase BMI vs all other (<25 BMI)** | | | **Decrease BMI vs all other (>=25 BMI)** | | |
|---|---|---|---|---|---|---|---|
| | | | **Type III Test** | | | **Type III Test** | |
| **Model** | **Predictor** | **QIC** | **W χ2** | **p** | **QIC** | **W χ2** | **p** |
| All predictors (N=673/743) | | 838.478 | | | 871.439 | | |
| | Gender | | 0.010 | 0.921 | | **4.132** | **0.042** |
| | Ethnicity | | 0.131 | 0.718 | | 2.434 | 0.119 |
| | Baseline BMI | | **8.589** | **0.003** | | 0.842 | 0.359 |
| | Age | | 2.782 | 0.095 | | **4.286** | **0.038** |
| | Occupation and work from home | | 4.436 | 0.109 | | 2.555 | 0.279 |
| | Socioeconomic score | | 4.434 | 0.218 | | 1.976 | 0.577 |
| | Living conditions | | 1.567 | 0.457 | | 2.749 | 0.253 |
| | Isolation status | | 0.279 | 0.597 | | 0.241 | 0.624 |
| | Quality of Life | | 0.407 | 0.523 | | 1.167 | 0.280 |
| | HFSS snacks intake | | 1.073 | 0.300 | | **7.113** | **0.008** |
| | HFSS meals intake | | 1.729 | 0.189 | | 0.132 | 0.716 |
| | Fruit and vegetables intake | | **5.628** | **0.018** | | **5.052** | **0.025** |
| | ΔHFSS snacks change score | | 1.881 | 0.170 | | 1.272 | 0.259 |
| | ΔHFSS meals change score | | 2.430 | 0.119 | | 0.581 | 0.446 |
| | ΔFruit and vegetables change score | | 0.100 | 0.752 | | 0.007 | 0.932 |
| | Physical activity | | 0.047 | 0.828 | | 0.015 | 0.904 |
| | Alcohol consumption | | 0.397 | 0.528 | | **12.353** | **<0.001** |
| | Smoking status | | <0.001 | 0.999 | | 1.709 | 0.191 |

*Models also included Time as a covariate. Type III tests are shown only. In the 'Increase BMI vs all other' GEE model, the baseline BMI continuous variable was converted to a categorical variable, due to violating the linearity of logit assumption. There were no material changes in significance of main effects. QIC is a relative, 'lower is better' measure of goodness of fit. W $\chi 2$: Wald Chi-square.*
